# Emergence of a *Pseudomonas aeruginosa* Hypermutator Strain During the Course of Ventilator-Associated Pneumonia

**DOI:** 10.1101/2022.05.19.22275029

**Authors:** Sophie Nozick, Egon A. Ozer, Rachel Medernach, Rebecca Kumar, Jori O. Mills, Richard G. Wunderink, Chao Qi, Alan R. Hauser

## Abstract

Hypermutator lineages of *Pseudomonas aeruginosa* arise frequently during the years of lung infection seen in patients with cystic fibrosis and bronchiectasis but are rare in the absence of structural lung disease. Since the onset of the COVID-19 pandemic, large numbers of patients have remained mechanically ventilated for extended periods of time. These patients are prone to acquire bacterial pathogens that persist for many weeks and have the opportunity to evolve within the pulmonary environment. However, little is known about what types of adaptations occur in these bacteria and whether these adaptations mimic those described in chronic infections. We describe a COVID-19 patient with a secondary *Pseudomonas aeruginosa* lung infection in which the causative bacterium persisted for >90 days. During the course of this infection, a hypermutator lineage of *P. aeruginosa* emerged and co-existed with a non-hypermutator lineage. Compared to the parental lineage, the hypermutator lineage evolved to be more extensively resistant to antibiotics, to change its type III secretion profile, and to grow more slowly. Genomic analyses of the hypermutator lineage identified numerous mutations, including in the mismatch repair gene *mutL* and other genes frequently mutated in individuals with cystic fibrosis. Together, these findings demonstrate that hypermutator phenotypes can emerge when clearance of *P. aeruginosa* fails to occur in typically acute infections such as ventilator-associated pneumonia and suggest that hypermutator lineages can affect patient treatments and outcomes.

## INTRODUCTION

Many bacterial pathogens experience substantial stress as they transition from their environmental reservoirs to the human body in the process of causing an infection. The immune system, intrinsic defenses, and altered nutrient availability together create an inhospitable environment to which the pathogen must adapt. Added to this are medically administered therapies such as antibiotics designed to eradicate the pathogen. To persist, the pathogen must adapt by the action of pre-programmed sensing systems or the acquisition of mutations. Adaptation to the human host has been most thoroughly studied in the context of chronic infections lasting for years. The role of adaptation in acute or subacute infections remains unclear.

Long-term respiratory infections with the bacterium *Pseudomonas aeruginosa* in individuals with cystic fibrosis (CF) have served as a model for the study of bacterial adaptation. These patients tend to remain infected with a single infecting lineage of *P. aeruginosa* for decades. Well characterized adaptations in this context include the emergence of a mucoid colony morphology, antibiotic resistance, loss of type III secretion, loss of motility, quorum sensing defects, and auxotrophy (1). Interestingly, it appears that the emergence of these traits is accelerated by the presence of a “hypermutator” phenotype in which an increased spontaneous mutation rate occurs following defects in DNA repair genes. Such hypermutator strains become fixed in populations exposed to new conditions in which the rapid emergence of mutations provides an adaptive advantage (2). In *P. aeruginosa*, hypermutability is most often a consequence of disruptions of mismatch repair genes, such as *mutS, mutL*, and *uvrD* (also called *mutU*) (3).

Two especially well studied phenotypes associated with hypermutator strains of *P. aeruginosa* in CF are the emergence of antibiotic resistance and altered pathogenicity. In general, hypermutator strains have a higher prevalence of resistance to antibiotics than non-mutators (4-6). Indeed, hypermutator strains acquire resistance to antibiotics more quickly than non-mutator strains (6) and even in susceptible strains resistance tends to emerge rapidly upon exposure to antibiotics (7, 8). Similarly, inactivation of the mismatch repair system of *P. aeruginosa* reduced *in vitro* fitness and attenuated virulence in acute infections. Hypermutator isolates recovered from CF patients demonstrated defective type III secretion (a major virulence factor of *P. aeruginosa* (9)) (10) and quorum sensing (11). Together, these observations indicate that the emergence of a hypermutator phenotype can have profound effects on antibiotic resistance and pathogenicity.

Hypermutator strains of *P. aeruginosa* are common in CF and other chronic infections associated with structural lung disease (4, 12-14). In contrast, these strains are relatively rare in acute infections, such as ventilator-associated pneumonia, in the absence of pre-existing structural lung abnormalities (4, 15). This has led to speculation that hypermutators are selected against in acute infections (4). However, the COVID-19 epidemic has led to large population of patients undergoing prolonged mechanical ventilation (16); these patients are at risk for *P. aeruginosa* ventilator-associated pneumonia or carriage that persists for weeks. It remains unclear whether these conditions allow for the emergence of hypermutators and whether such emergence has the potential to affect the treatment and outcome of pneumonia.

Here, we present a non-CF patient with COVID-19 acute respiratory distress syndrome (ARDS) complicated by a protracted case of ventilator-associated pneumonia during which *P. aeruginosa* developed a hypermutator phenotype. Over the course of the infection, a large number of mutations occurred in the *P. aeruginosa* lineage, including some that were associated with marked changes in antibiotic resistance and pathogenicity. This case demonstrates that hypermutators can evolve in the absence of chronic infections and have the potential to affect patient outcomes.

## MATERIALS AND METHODS

### Patient enrollment and isolate collection

Clinical information was obtained from the medical records of the hospitals to which he was admitted. Clinical bacterial isolates were obtained from the microbiology laboratory at the second hospital following collection during routine clinical care. Each isolate was archived at −80ºC. This study was approved by the Northwestern University Institutional Review Board (IRB). For certain experiments, *P. aeruginosa* strains PA14, PA14*exoU*, and PAO1 were used as controls.

### Whole-genome sequencing

Isolates were grown overnight in Luria Bertani (LB) broth with shaking at 37°C. DNA extraction was performed using a Maxwell 16 Instrument with Cell DNA purification kits (Promega Corporation, Madison, WI) according to the manufacturer instructions. For short-read sequencing, libraries were prepared using the Illumina Nextera XT kit and sequenced using an Illumina NextSeq (150-bp paired-end reads, high-output) platform (Illumina, Inc., San Diego, CA). Sequences were trimmed using Trimmomatic v0.32 (17) and then *de novo* assembly was performed with SPAdes 3.9.1. Contigs were removed if they were shorter than 200□bp or had a mean fold coverage of <5x per base. For strain L00-a, additional long-read sequencing was performed using the Nanopore MinION platform, and hybrid assembly with short-read Illumina sequences was performed. Briefly, long-read sequencing libraries were prepared from unsheared genomic DNA using ligation sequencing kit SQK-LSK109 (Oxford Nanopore, UK) and sequenced on the MinION platform using a FLO-MIN106 flow cell. Guppy v3.4.5 was used to base call reads with the R9.4.1 high-accuracy model and to perform read quality filtering based on Q scores, demultiplexing, and barcode trimming. Assembly of Nanopore reads was performed using the Trycycler v0.5.0 pipeline (18) as follows: Raw nanopore reads were first filtered using Filtlong v0.2.1 (https://github.com/rrwick/Filtlong) to remove reads shorter than 1000 bases and the lowest quality 5% of the reads. Reads were then subsampled into 12 subsets using the Trycycler “subsample” function. Four read subsets were assembled using Flye v2.8.3 (19) or (20) or minimap2 v2.21, miniasm v0.3, minipolish v0.1.3 via the “miniasm_and_minipolish.sh” script (https://github.com/rrwick/Minipolish/blob/main/miniasm_and_minipolish.sh). Clustering, reconciliation, circularization, multiple sequence alignment, and consensus sequence generation from the resulting 12 assemblies were performed using Trycycler. The resulting consensus assembly was polished with the nanopore long reads and medaka v1.4.3 using the “r941_min_hac_g507” model. Illumina reads were aligned to the assembly using BWA v0.7.17 (21), and assembly errors were corrected using Pilon v1.23 (22) with a minimum depth setting of 0.1. Serial read alignment and Pilon correction were performed sequentially until no further assembly corrections were generated. Polypolish v0.4.3 (23) was used to further correct the assembly using the Illumina reads. Custom software (Pilon Tools v0.1; https://github.com/egonozer/pilon_tools) was used to identify, manually assess, and correct any residual homopolymer assembly errors. Sequences were deposited in the National Center for Biotechnology Information (NCBI) database (see Supplemental Table 1 for accession numbers).

### Single nucleotide variant identification and phylogenetic analysis

Illumina short reads for each isolate were aligned to the genome sequence of strain L00-a using bwa v 0.7.15 (21). Single nucleotide variants relative to the reference were identified using bcftools v1.9 skipping bases with base quality lower than 25, alignment quality less than 30, and using a haploid model. Variants were further filtered as previously described (24) using the bcftools_filter software (https://github.com/egonozer/bcftools_filter) to remove variants with single nucleotide variant (SNV) quality scores less than 200, read consensuses less than 75%, read depths less than 5, read numbers in each direction less than 1, or locations within repetitive regions (as defined by blast alignment of the reference genome sequence against itself). The resulting multiple sequence alignments were filtered to include only variant positions with a defined base in each of the 22 isolates, i.e., the 100% core genome. A maximum likelihood phylogenetic tree was generated from the core genome alignment with IQ-TREE v1.6.1 using the ModelFinder function to estimate the best-fit nucleotide substitution model by means of Bayesian information criterion (BIC) (25, 26). Tree topology was assessed both with the Shimodaira-Hsegawa approximate likelihood ratio test (SH-aLRT) and with the ultrafast booststrap (UFboot) with 1,000 replicates each (27, 28).

### Rifampin-resistance assays

Mutation rates were quantified using assays to measure the emergence of resistance to rifampin, as described by Oliver and colleagues (12). Briefly, a bacterial colony was resuspended in 20 ml of Mueller-Hinton (MH) broth and grown at 37°C overnight. Bacterial cells were then collected by centrifugation at 3,000 x g for 5 min. Bacteria were resuspended in 1 ml of MH broth, and either 10 μl or 100 μl aliquots of serially diluted samples were plated onto MH agar plates with and without rifampicin supplementation (300 mg/ml). It was demonstrated that each isolate was initially susceptible to this concentration of rifampicin. The plates were incubated at 37°C for 24-48 hr, and the colonies were counted. All experiments were performed in triplicate and the mean value was recorded. A strain was designated a hypermutator when the corresponding mutation frequencies for rifampicin were at least 20-fold higher than those observed for the initially cultured isolate, L00-a, and reference strain PAO1 (12), and when the absolute resistance frequency was >2 × 10^−7^, as previously described (11).

### Bacterial growth assays

A single bacterial colony was inoculated into 5 ml of LB broth and grown overnight at 37°C at 250 rpm. The following morning, the bacteria was sub-cultured 1:100 in fresh LB for 3 hours and then adjusted to an optical density at 600 nm (OD_600_) of 0.1 in LB broth. A total of 200 μl of each culture was added to a 96-well clear, flat-bottom plate (Corning, Corning, NY). The plate was incubated at 37°C at 250 rpm, and bacterial growth was quantified using a BioTek Synergy H1 instrument to measure OD_600_ every 30 min over 24 hr. Each isolate was tested in triplicate.

### Antibiotic susceptibility assays

A sample from a cryopreserved stock was streaked on an LB agar plate and grown overnight at 37°C at 250 rpm. Bacterial colonies were resuspended in sterile water and then standardized to an OD_600_ of 0.08-0.1. The bacterial sample was then diluted in MH broth and distributed into pre-manufactured Sensititre GNX3F plates (Thermo Fisher, Scientific U.K.) The plates were incubated for 20 hr and were manually read. All isolates grew in the wells lacking antibiotics. Clinical and Laboratory Standards Institute (CLSI) 2021 breakpoints for *P. aeruginosa* were used. Assays were performed twice on each isolate. If MIC results differed by more than one dilution, the assay was performed a third time, and the majority result was reported.

### Immunoblot analyses

For detection of ExoU secretion, *P. aeruginosa* strains were grown in LB broth overnight at 37°C at 250 rpm. The bacteria were sub-cultured in LB broth with and without MgCl_2_ (20 mM) and EGTA (5 mM) for 3 hr at 37°C at 250 rpm. Bacterial supernatants were obtained by centrifugation at 2500 x g for 10 minutes at room temperature. The supernatant was passed through a 0.22 μm filter and resuspended in trichloroacetic acid overnight at 4°C with slow shaking. The precipitated proteins were collected by centrifugation at 8,650 x g for 45 min at 4°C, washed two times with acetone, and then re-suspended in Laemmli Sample Buffer (Bio-Rad, Hercules, CA). The sample was heated to 80°C for 10 min and 20 μL of each sample was electrophoresed through a 4-15% precast polyacrylamide gel (Bio-Rad, Hercules, CA). Proteins were then electrotransferred onto a nitrocellulose membrane (Schleicher and Schuell). The membrane was blocked with 5% milk powder (Boston Bioproducts, Ashland, MA) in 1x tris-buffered saline (Bio-Rad, Hercules, CA) with 0.5% Tween-20 (TBS-T; National Diagnostics, Atlanta, GA) for 1 hr at room temperature and then exposed to a 1:2,000 concentration of polyclonal anti-ExoU antibodies overnight at 4°C on a slow rocker (29). Polyclonal ExoU antiserum was prepared as described elsewhere (30). The membrane was washed three times with 1x TBS-T at room temperature and then exposed to a goat anti-rabbit secondary antibody (LI-COR Biosciences, Lincoln, NE) at a 1:5,000 concentration in 20 mL of 5% milk buffer in 1x TBS-T for 1 hr at room temperature. Membranes were incubated in TBS-T and visualized using a LI-COR Biosciences Odyssey Fc Imaging System.

## RESULTS

### Case presentation

The patient was a previously healthy man in his 50s with no prior past medical history who admitted to a local hospital with COVID-19 pneumonia (**Fig. 1**). He required mechanical ventilation on hospital day 8, at which time a respiratory culture grew *P. aeruginosa* and *Klebsiella aerogenes* and he was diagnosed with a secondary *P. aeruginosa* pneumonia. He developed ARDS and on hospital day 10 required veno-venous extracorporeal membrane oxygenation (VV ECMO) support. The following day, his respiratory cultures grew *P. aeruginosa*. Six days later a bronchoscopy was performed, and bronchoalveolar lavage (BAL) fluid again grew *P. aeruginosa*. On hospital day 20, blood cultures grew *P. aeruginosa*. On hospital days 22 and 31, BAL fluid grew *P. aeruginosa* and *Serratia marcescens*, and on hospital day 35 *P. aeruginosa*. The patient was transferred to a second medical center on hospital day 45 for consideration for lung transplantation. On that day and two days later, BAL fluid was obtained and subsequently grew *P. aeruginosa*. CT imaging of the lungs was notable for a right middle lobe fluid collection concerning for a possible abscess. The patient was treated with piperacillin-tazobactam. His course was also complicated by a tension hemopneumothorax secondary to traumatic chest tube placement. On hospital day 49, the patient developed septic shock with worsening lung function. His antibiotics were changed to ceftazidime/avibactam plus ciprofloxacin, and he was started on veno-arterial (VA) ECMO. His condition stabilized, and he was transitioned to VV ECMO. Two days later his antibiotics were changed to cefepime, metronidazole, and inhaled amikacin, which were stopped after completion of a 2-week course. On hospital day 75, the patient developed a worsening pressor requirement, and blood cultures and BAL fluid again grew *P. aeruginosa*. Cefepime was started but was changed to ceftazidime/avibactam two days later. Blood cultures drawn on hospital day 77 and 78 remained positive for *P. aeruginosa*, and on hospital day 79 antibiotics were transitioned to ciprofloxacin plus polymyxin B based on susceptibility results. On hospital days 82, 88, 91, and 100, BAL fluid was obtained that all subsequently grew *P. aeruginosa*. Unfortunately, the patient was ultimately determined to be a poor transplantation candidate, and withdrawal of supportive measures ensued. The patient succumbed to his illness on hospital day 107.

**Figure 1.**
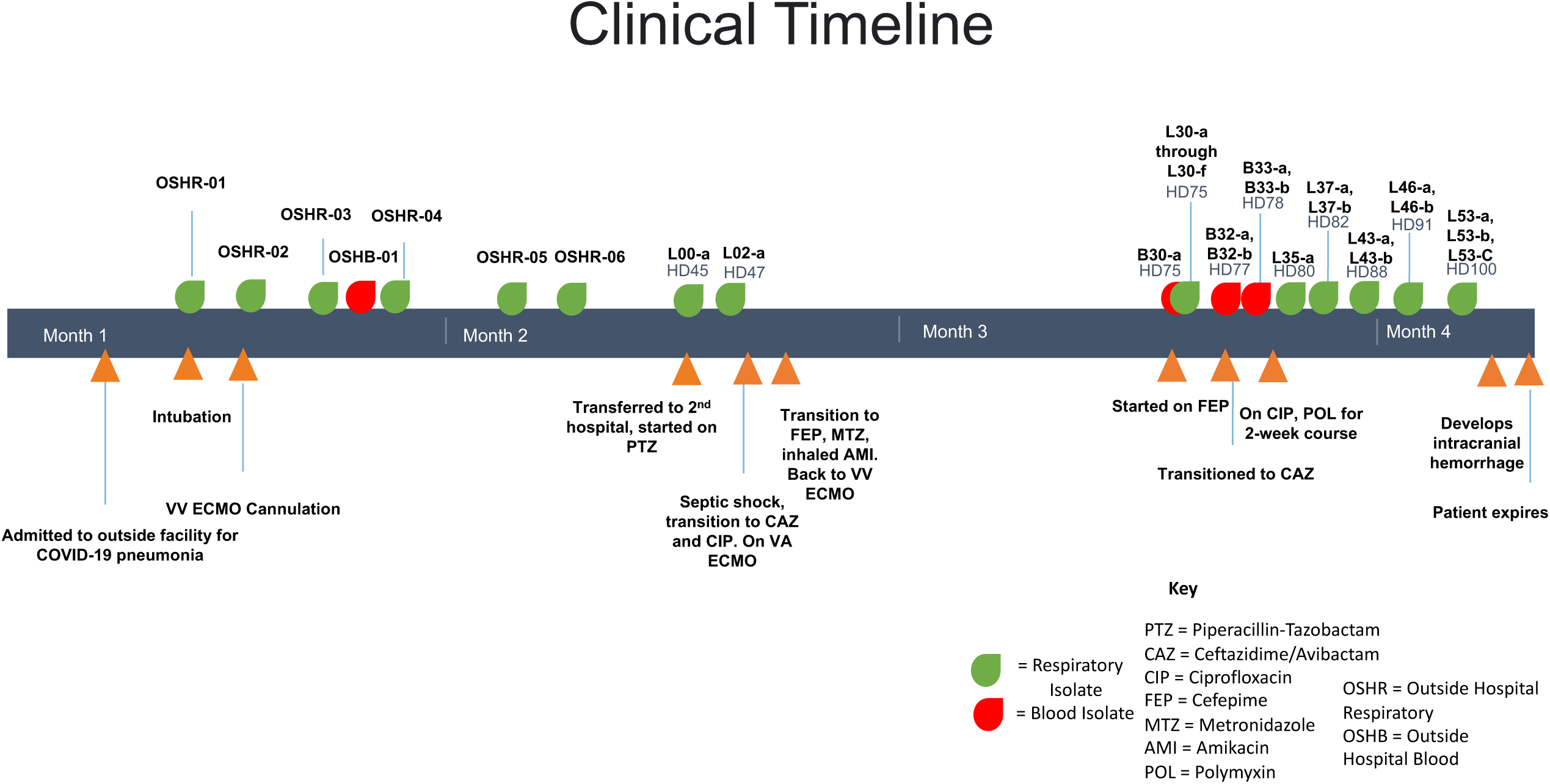
The clinical course of the patient indicating when medical events occurred and when each *P. aeruginosa* isolate was obtained.

### Phylogenetic relationships of *P. aeruginosa* isolates

A total of 23 *P. aeruginosa* isolates were from the patient were available for analysis. The complete genome sequence of the first available *P. aeruginosa* isolate (L00-a obtained on hospital day 45) was obtained using hybrid assembly of both the long-read Nanopore and short-read Illumina platforms. The remaining 22 isolates were sequenced using the Illumina platform and were aligned to the L00-a genome to identify SNVs that evolved during the infection. Phylogenetic analysis of the 23 sequences demonstrated that all were of the same lineage (i.e., all isolates were descendants of the initial infecting bacterium) (**Fig. 2A**). However, two distinct patterns were readily apparent. Many of the isolates, including the isolate collected 2 days after L00-a, possessed relatively few SNVs (5-21) compared to L00-a (**Fig. 2B**). Isolates from this group were cultured throughout the patient’s hospitalization. However, beginning on hospital day 75 many isolates were obtained that had acquired relatively large numbers of SNVs (>100). Isolates such as these were also present throughout the remaining course of infection, co-existed with the first lineage, and were cultured from both respiratory and blood samples.

**Figure 2.**
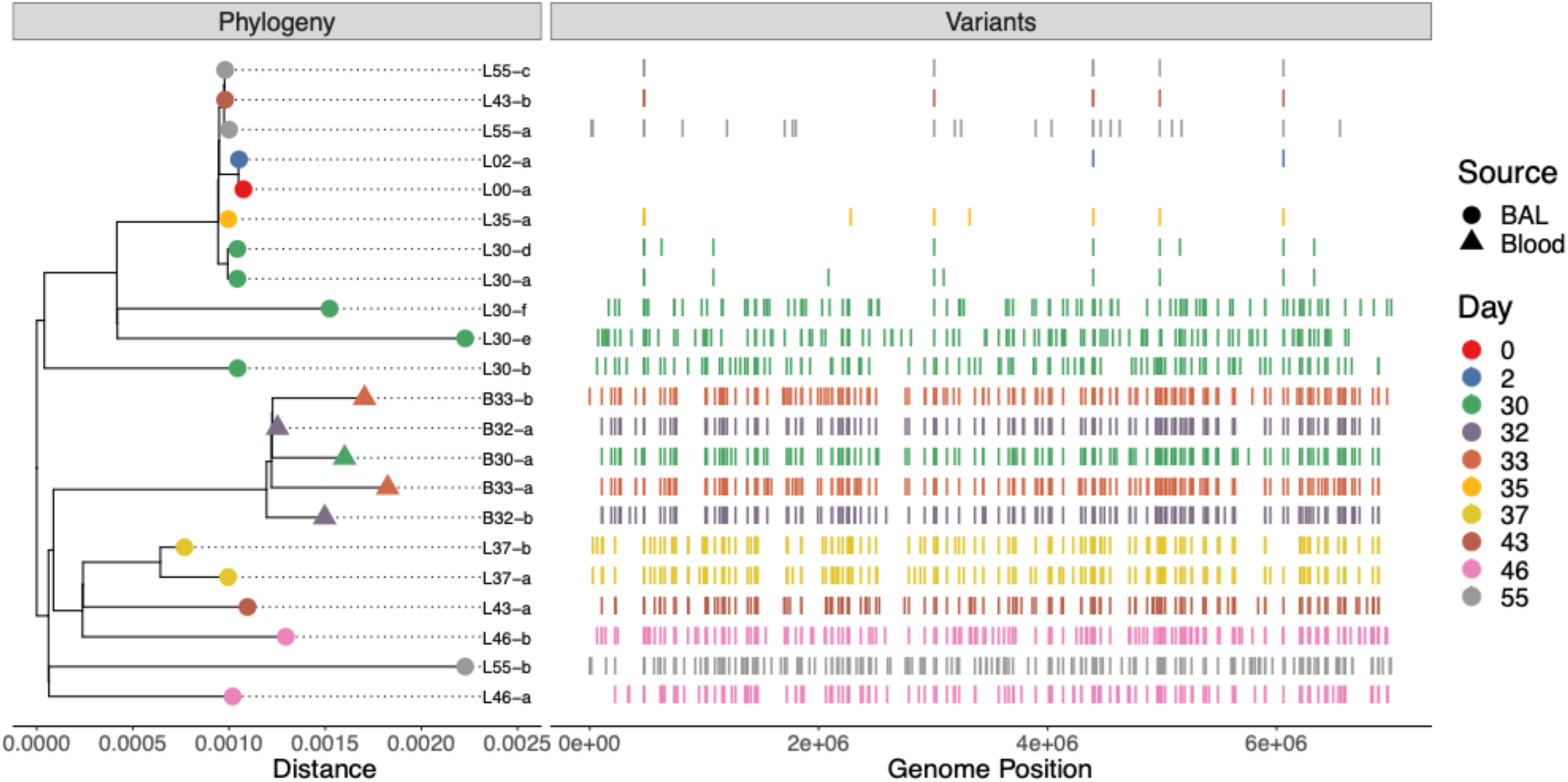
Whole genome sequencing of *P. aeruginosa* isolates. On the left, a phylogenetic tree is shown. On the right, the locations of SNVs across the genome are indicated.

### Hypermutator analyses

The subpopulation of *P. aeruginosa* isolates containing numerous mutations that evolved over the relatively short time of the patient’s hospitalization suggested a hypermutator strain (11). Indeed, a SNV yielding a nonsynonymous single-amino-acid substitution in the mismatch repair gene *mutL* was noted to be present in each of the putative hypermutator isolates but in none of the other isolates (Supplemental Table 1). To further explore the significance of this mutation, we performed assays to measure the emergence of resistance to rifampin using 3 representative isolates with the *mutL* SNV and 3 isolates without the *mutL* SNV. Since resistance to rifampin requires a single mutation, spontaneous emergence of rifampin resistance can be used as a phenotypic surrogate to measure mutation rate (31). Broth cultures of isolates were grown in the absence of rifampin and then plated on agar supplemented with or without rifampin to quantify the rate at which rifampin-resistant bacteria evolved in strains with wild-type and mutated *mutL*. The three isolates containing *mutL* mutations evolved rifampin resistance at approximately 100-fold greater frequency than the isolates containing intact *mutL* and the control strain PAO1 (**Fig. 3**). These findings confirm that a *P. aeruginosa* hypermutator lineage evolved during the course of the patient’s infection.

**Figure 3.**
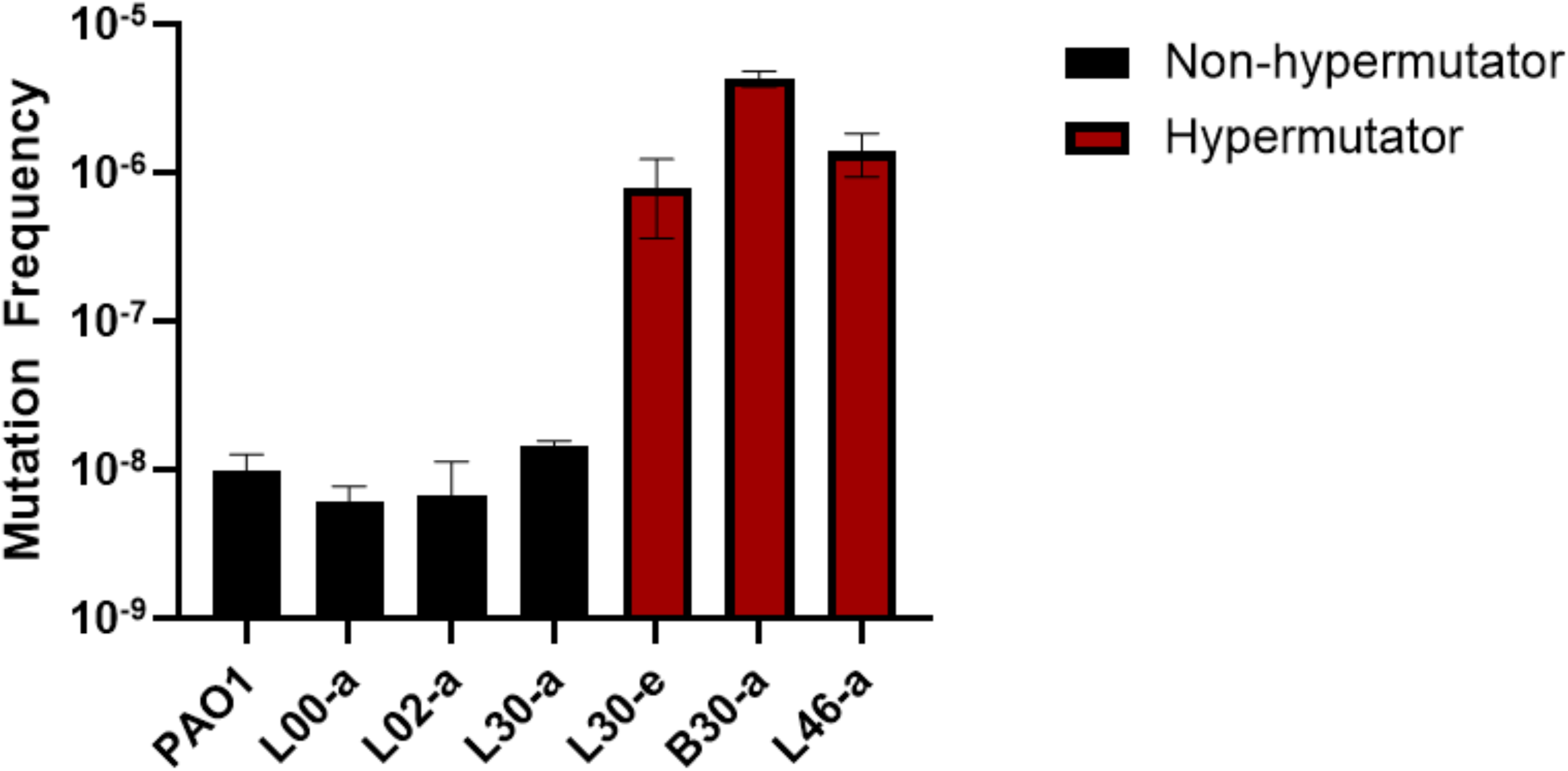
Emergence of resistance to rifampin in representative hypermutator and non-hypermutator isolates. Strain PAO1 is a reference non-hypermutator *P. aeruginosa* strain. Each experiment was performed 2-3 times.

### Antibiotic resistance

One of the fitness pressures that may select for the emergence of hypermutator strains is the inhibitory activity of antibiotics. Hypermutator strains acquire resistance to antibiotics more quickly than conventional strains, which may allow them to better persist in the face of antimicrobial therapy. As mentioned, this patient was heavily exposed to antipseudomonal antibiotics, including piperacillin-tazobactam, ceftazidime-avibactam, ciprofloxacin, cefepime, polymyxin B, and inhaled amikacin. We therefore examined the antibiotic susceptibilities of the isolates using broth dilution assays. The earliest available *P. aeruginosa* isolate from this patient (L00-a) was susceptible to each of the tested antibiotics (**Fig. 4**). However, antibiotic resistance subsequently developed in both hypermutator and non-hypermutator lineages. An isolate cultured only two days later (L02-a) had acquired resistance to imipenem and doripenem and intermediate susceptibility to meropenem. Non-hypermutator isolates collected on or after hospital day 75 were largely resistant to carbapenems, cefepime, and ceftazidime (**Fig. 4**), while hypermutator isolates were resistant to these antibiotics plus gentamicin and aztreonam. In addition, hypermutator isolates were only intermediately susceptible to piperacillin-tazobactam and amikacin. Beginning on hospital day 82 hypermutator isolates were resistant to ciprofloxacin and levofloxacin. Although the non-hypermutator lineage presumably predated the hypermutator lineage and therefore was exposed to antibiotics for a longer time, it developed slightly less non-susceptibility (resistance or intermediate susceptibility) than the hypermutator lineage (9 of the 14 vs. 11 of the 14 tested antibiotics). These findings are consistent with reports showing that hypermutator strains are prone to develop antibiotic resistance in chronic infections (13).

**Figure 4.**
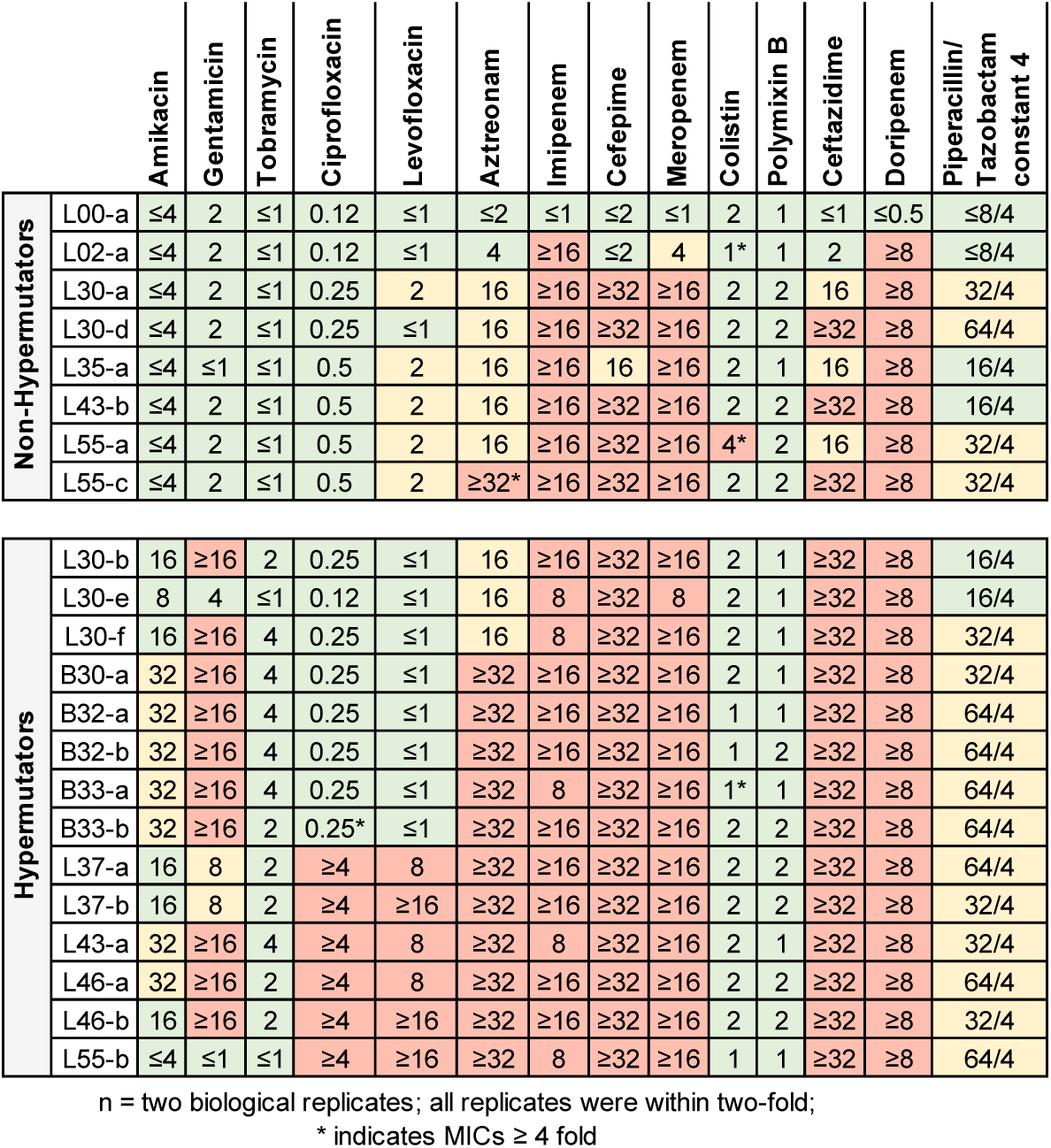
The minimum inhibitory concentrations (MICs) of *P. aeruginosa* isolates to antibiotics. Susceptible MICs are shown in green, intermediately susceptible MICs in yellow, and resistant MICs in red.

An analysis of the genomes of the *P. aeruginosa* isolates provided mechanistic explanations for some of the phenotypic antimicrobial resistance patterns. In both hypermutator and non-hypermutator lineages, intermediate susceptibility to aztreonam and resistance to meropenem, cefepime, and ceftazidime were associated with three changes that appeared simultaneously in hospital day 77 isolates: (1) a glycine-to-tryptophan substitution at amino acid 101 of MexR, (2) an arginine-to-cysteine substitution at amino acid 620 of MexB, and a substitution causing a premature stop codon in the *oprD* gene. Since these changes occurred in both lineages, they likely happened prior to the emergence of the hypermutator lineage. MexR is a repressor of the MexAB-OprM efflux pump, and its disruption could lead to overexpression of MexAB-OprM, which has been linked to beta-lactam resistance (32). Likewise, the substitution in MexB has the potential to enhance efflux of beta-lactams. Disruption of the OprD outer membrane porin contributes to high-level resistance to carbapenems (32). The hospital day 77 hypermutator isolates (but not the non-hypermutator lineage) also contained an arginine-to-histidine substitution at residue 504 of penicillin-binding protein 3, which may be responsible for the elevated MICs to aztreonam observed in this lineage (32). On hospital day 84, the hypermutator lineage acquired a threonine-to-isoleucine substitution in subunit A of DNA gyrase, which was associated with the development of resistance to ciprofloxacin and levofloxacin. These findings demonstrate how increased numbers of mutations at the genomic level may lead to phenotypic antibiotic resistance in hypermutator strains.

### Virulence

Because hypermutator strains have also been linked to changes in pathogenicity (33), we performed immunoblot analyses on several of the isolates to examine type III secretion of ExoU, a major virulence determinant of *P. aeruginosa*. We tested hypermutator isolates from both the lungs (L30-b, L30-e, L30-f) and the blood (B30-a, B32-a, B32-b, B33-a, B33-b). Immunoblots demonstrated that all these isolates except one (the hypermutator L30-f) secreted ExoU (**Fig. 5**). These findings demonstrate that defective type III secretion evolved in the hypermutator lineage, although it may be a relatively rare occurrence.

**Figure 5.**
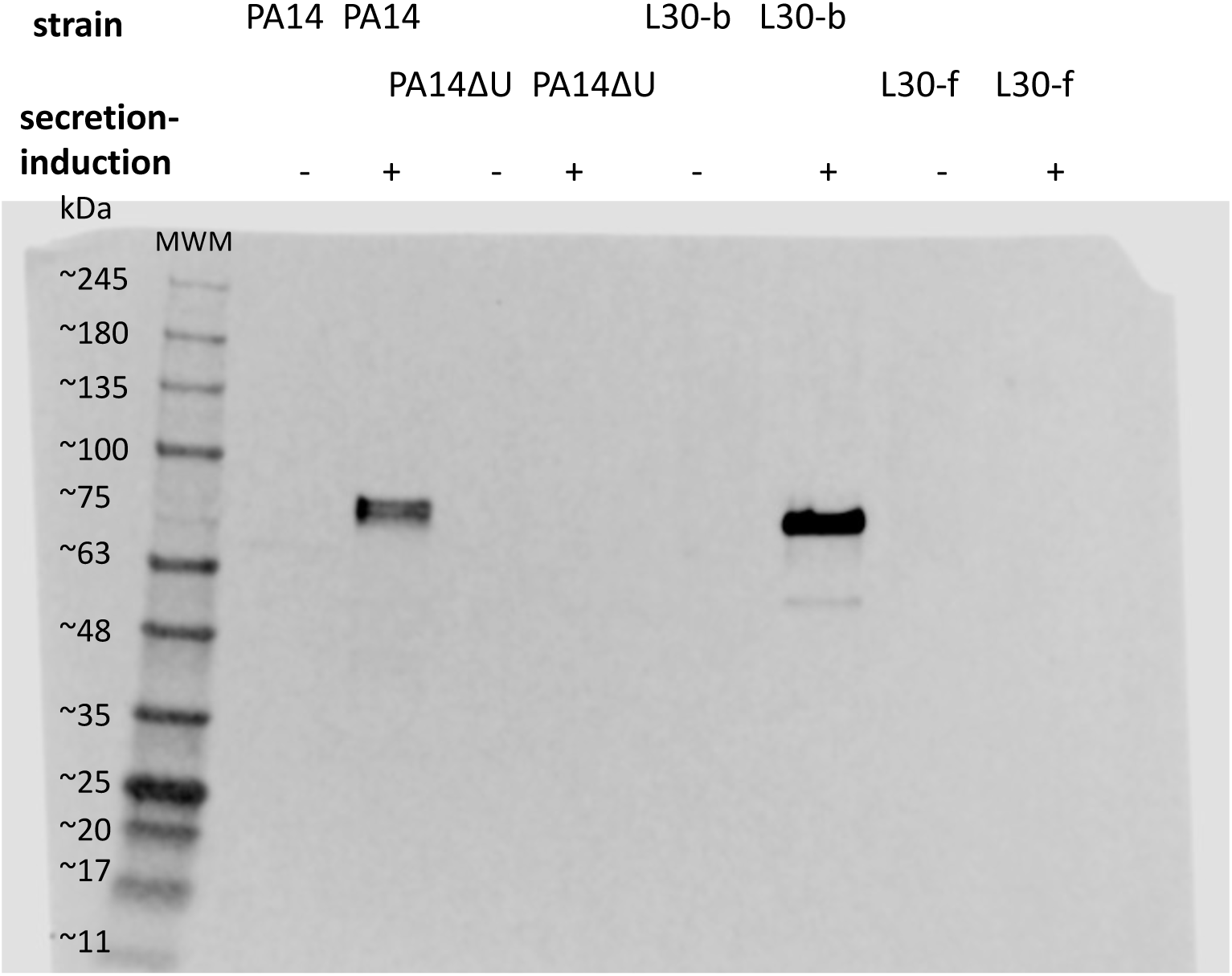
Immunoblot for detection of secreted ExoU by representative *P. aeruginosa* antibodies. Bacterial culture supernatants were precipitated, concentrated, and tested for the presence of ExoU using polyclonal anti-ExoU antibodies. PA14 is an ExoU-secreting reference strain, and PA14ΔU is the same strain containing a deletion in the *exoU* gene.

### Growth rate

Hypermutator strains contain large numbers of mutations, so one would anticipate that their growth rates may be affected. We therefore measured the growth rates of the 23 isolates in rich (LB) medium. Growth rates differed substantially among the different isolates, and all isolates grew somewhat more slowly than the control strain PAO1 (**Fig. 6**). The slowest growing isolate, L55-b, was a hypermutator strain, and several other hypermutator isolates required more time to reach stationary growth phase than the non-hypermutator isolates. Compared to the non-hypermutator isolates, the hypermutator isolates showed a trend towards slower growth that was not statistically significant (two sample t-test, p = 0.096).

**Figure 6.**
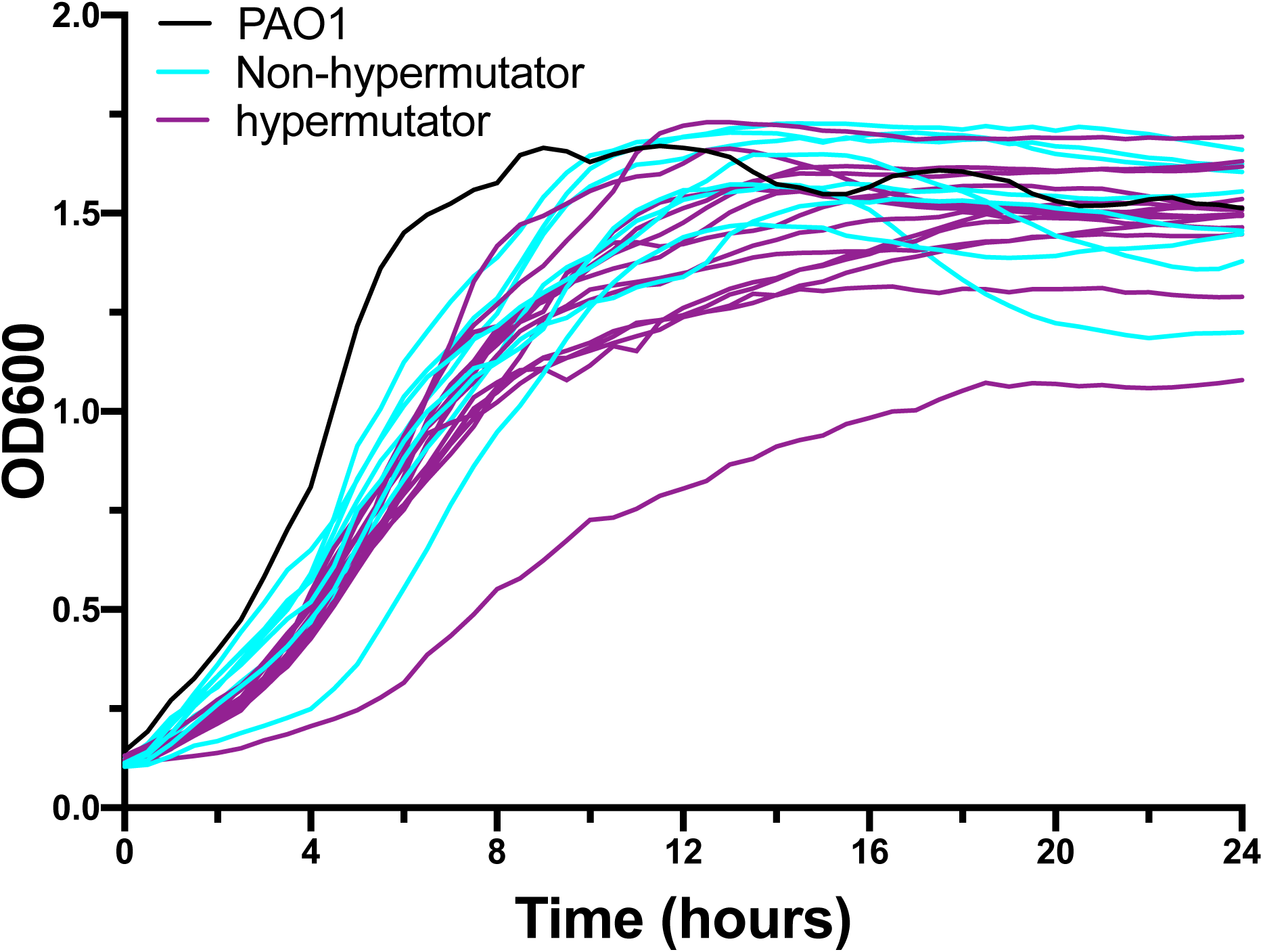
Growth curves of *P. aeruginosa* isolates. PAO1 is a non-hypermutator reference *P. aeruginosa* strain.

### Genes containing nonsynonymous mutations

We next examined the genes that most commonly contained non-synonymous mutations in the collection of *P. aeruginosa* isolates. We reasoned that commonly observed mutations may reflect adaptations that provide a fitness advantage to *P. aeruginosa* in the face of the pulmonary environment or antibiotic therapy. Most of the hypermutator and non-hypermutator isolates contained a mutation in the *lasR* gene, which encodes a regulator of the *las* quorum-sensing system (**Table 1**). Other commonly mutated genes included *mexR* (regulator of the MexAB-OprM multidrug efflux pump), *oprD* (porin that allows entry of carbapenems), and *pilB* (biogenesis of type 4 pili). A complete list of mutations can be found in Supplemental Table 1. These findings demonstrate that some mutations were commonly observed in all isolates, whereas others were seen only in hypermutator isolates.

**Table 1.** Genes in which nonsynonymous mutations were most commonly observed in the patient’s P. aeruginosa isolates.

| Product | Amino acid change | # of mutants relative to first iso | # of Blood Isolates | # of Hypermutators | # of Non-Hypermutators |
| --- | --- | --- | --- | --- | --- |
| transcriptional regulator LasR [PA1430] | M25K | 21 | 5 | 14 | 7 |
| multidrug resistance operon repressor MexR [PA0424] | G101W | 20 | 5 | 14 | 6 |
| Resistance-Nodulation-Cell Division (RND) multidrug efflux transporter MexB [PA0426] | R620C | 20 | 5 | 14 | 6 |
| sigma factor PvdS [PA2426] | V71A | 20 | 5 | 14 | 6 |
| Basic amino acid, basic peptide and imipenem outer membrane porin OprD precursor [PA0958] | W339* | 20 | 5 | 14 | 6 |
| hypothetical protein [PA4048] | A133V | 14 | 5 | 14 | 0 |
| hypothetical protein [PA3722] | A69G | 14 | 5 | 14 | 0 |
| D-serine dehydratase [PA3357] | F29L | 14 | 5 | 14 | 0 |
| putative two-component sensor [PA3044] | R466W | 14 | 5 | 14 | 0 |
| soluble pyridine nucleotide transhydrogenase [PA2991] | I143V | 14 | 5 | 14 | 0 |
| nitrate transporter [PA1783] | G381D | 14 | 5 | 14 | 0 |
| hypothetical protein [PA0830] | Y244H | 14 | 5 | 14 | 0 |
| hypothetical protein | H114R | 14 | 5 | 14 | 0 |
| putative transcriptional regulator [PA0791] | R108W | 14 | 5 | 14 | 0 |
| penicillin-binding protein 3 [PA4418] | R504H | 14 | 5 | 14 | 0 |
| type 4 fimbrial biogenesis protein PilB [PA4526] | V358M | 14 | 5 | 14 | 0 |
| ferric iron-binding periplasmic protein HitA [PA4687] | G55S | 14 | 5 | 14 | 0 |
| DNA mismatch repair protein MutL [PA4946] | H472L | 14 | 5 | 14 | 0 |
| putative binding protein component of ABC transporter [PA5076] | T104A | 14 | 5 | 14 | 0 |
| hypothetical protein | I62V | 14 | 5 | 14 | 0 |
| porphobilinogen deaminase [PA5260] | D293G | 14 | 5 | 14 | 0 |
| hypothetical protein [PA3712] | L106P | 13 | 5 | 13 | 0 |
| gluconate dehydrogenase [PA2265] | G39S | 13 | 5 | 13 | 0 |
| 3-hydroxybutyrate dehydrogenase [PA2003] | G89S | 13 | 5 | 13 | 0 |
| hypothetical protein [PA1789] | R127Q | 13 | 5 | 13 | 0 |
| putative transporter [PA1419] | V94I | 13 | 5 | 13 | 0 |
| putative siderophore receptor [PA1365] | Q511R | 13 | 5 | 13 | 0 |
| hypothetical protein [PA1045] | S460G | 13 | 5 | 13 | 0 |
| TadZ [PA4303] | A271T | 13 | 5 | 13 | 0 |
| hypothetical protein [PA4318] | G240D | 13 | 5 | 13 | 0 |

## DISCUSSION

Hypermutator strains have been associated with increased antibiotic resistance and altered virulence phenotypes in *P. aeruginosa* lineages causing chronic infections, especially those in CF. In the case of ventilator-associated pneumonia reported here, a *P. aeruginosa* hypermutator lineage emerged over weeks to months rather than years. Although both the non-hypermutator and hypermutator lineages developed resistance to several antibiotics, resistance was more extensive in the hypermutator lineage. Likewise, one isolate of the hypermutator lineage lost the ability to secrete ExoU, a major *P. aeruginosa* virulence determinant. These results demonstrate that hypermutator strains can emerge over relatively short periods of time in individuals lacking pre-existing structural lung disease. In addition, these lineages have the potential to influence both antibiotic resistance and virulence.

While hypermutator strains of *P. aeruginosa* have been detected in 10—54% of individuals with CF (4), they are much less common in acute infections. For example, Oliver and colleagues did not detect hypermutator strains in *P. aeruginosa* isolates cultured from 50 patients with bacteremia and 25 patients with non-CF respiratory infections or colonization (12). Similarly, Gutierrez and colleagues tested 160 *P. aeruginosa* isolates from 103 non-CF patients and noted only 1 patient with a hypermutator strain (15). More recently, Torrens and colleague examined 723 *P. aeruginosa* isolates cultured from 402 patients in European intensive care units and found that only 2 were hypermutators (34). Even in CF patients, hypermutators tend to emerge after years of infection rather than subacutely; studies have found only 0-10% of newly infected CF patients harbor hypermutator strains whereas 24-73% of chronically infected CF patients harbor these strains (5, 11, 35). Our patient did not have CF or other known structural lung disease. He did, however, require prolonged ventilatory support, which is a risk factor for secondary bacterial pneumonia. Despite repeated bouts of antibiotics and partial clinical resolution, he failed to achieve microbiological eradication. This prolonged persistence of *P. aeruginosa* likely provided the opportunity for the bacterium to evolve into the hypermutator phenotype. Among respiratory bacteria that cause acute pneumonia, *P. aeruginosa* is notable for its ability to persist even in the face of active antibiotics (36, 37). Our findings demonstrate that hypermutator strains of *P. aeruginosa* may indeed arise in the setting of acute or subacute infections in the absence of pre-existing structural lung disease and that selective pressures within the lungs may allow these lineages to persist for weeks.

*P. aeruginosa* is notorious for its ability to develop resistance to antibiotics during the course of therapy, which occurs in approximately 10% of patients (38, 39). As mentioned, hypermutator strains of *P. aeruginosa* are especially prone to do so (4-6). The patient reported here illustrates these phenomena. Resistance to some carbapenems was observed even prior to the emergence of the hypermutator lineage (**Fig. 4**). Further loss of susceptibility to other antibiotics was observed in the non-hypermutator lineage later in infection and involved high levels of resistance to cefepime and ceftazidime, and intermediate susceptibilities to aztreonam and levofloxacin. However, during this same time the hypermutator strain acquired additional high-level resistance to aztreonam, levofloxacin, ciprofloxacin, and gentamicin. For amikacin and piperacillin-tazobactam, many isolates showed MICs in the intermediate range. Because both hypermutator and non-hypermutator lineages co-existed in the same patient, these findings nicely illustrate the increased propensity of hypermutator strains to develop resistance to antibiotics. They also indicate that a single isolate may provide misleading information regarding the selection of appropriate antimicrobial therapy.

Our findings also demonstrate the ways in which hypermutation may lead to changes in virulence. One of the hypermutator isolates lost the ability to secrete ExoU, a type III secretion effector protein that has been associated with worse outcomes in *P. aeruginosa* infections both in experimental models and human infections (40, 41). Although examination of more patients and isolates is necessary, this observation suggests that the increased difficulty in treating antibiotic-resistant hypermutator infections may be offset by a decrease in the intrinsic virulence of these strains.

One interesting aspect of our findings is that several mutations in the *P. aeruginosa* isolates from our patient occur commonly in CF infections. For example, most of our isolates contained mutations in *lasR*, which are also frequently observed in individuals with CF (1). This suggests that the fitness pressure driving emergence of these mutations is not specific to CF but to antibiotic therapy or the pulmonary environment in general. A remaining question is how long *P. aeruginosa* must be exposed to these fitness pressure before these mutations emerge.

Together, these findings suggest that *P. aeruginosa* hypermutator strains may emerge during acute and subacute infections in the absence of pre-existing lung disease and have the potential to affect virulence as well as response to treatment. Additional studies are necessary to determine how commonly hypermutators arise following ventilator-associated pneumonia and other acute or subacute infections. If these findings are generalizable, they may suggest that treatment strategies (adequate source control; highly active and pharmacologically optimized empiric antibiotic regimens) for these infections should be designed for rapid eradication of *P. aeruginosa* before the organism can evolve to become more difficult to treat.

## Data Availability

All data produced in the present study are available upon reasonable request to the authors

## ACKNOWLEDGEMENTS

We gratefully acknowledge the contributions of the NU SCRIPT Study Investigators. The project was funded by the National Institutes of Health: U19AI135964 (to R.G.W.) and 01AI118257, K24AI04831, R21AI153953, and R21AI164254 (to A.R.H.).

