## Supplemental Table 1 for "Emergence of a *Pseudomonas aeruginosa* Hypermutator Strain During the Course of Ventilator-Associated Pneumonia"

**Supplemental Table 1. Mutations in *P. aeruginosa* isolates**

| position | ref_base | locus_id | product | strand | gene_positio | coodn_position | codon | L00-a | L02-a | L30-a | L30-b | L30-d | L30-e | L30-f | B30-a | B32-a | B32-b | B33-a | B33-b | L35-a | L37-a | L37-b | L43-a | L43-b | L46-a | L46-b | L55-a | L55-b | L55-c |
|---|---|---|---|---|---|---|---|---|---|---|---|---|---|---|---|---|---|---|---|---|---|---|---|---|---|---|---|---|---|
| 16469 | C | OPLDDDIH_00011 | hypothetical protein [PA0007] | + | 1501 | 1 | CGG | R | R | R | R | R | R | R | R | R | R | R | R | R | R | R | R | R | R | R | R | W | R |
| 18405 | G | OPLDDDIH_00012 | glycyl-tRNA synthetase beta chain [PA0008] | - | 385 | 1 | CCG | P | P | P | P | P | P | P | P | P | P | P | P | P | P | P | P | P | P | P | X | S | P |
| 78599 | A | OPLDDDIH_00067 | putative transcriptional regulator [PA0056] | - | 422 | 2 | CTG | L | L | L | L | L | P | L | L | L | L | L | L | L | L | L | L | L | L | L | L | L | L |
| 112294 | T | OPLDDDIH_00098 | hypothetical protein [PA0087] | + | 37 | 1 | TCG | S | S | S | S | S | S | S | S | S | S | S | S | S | S | S | S | S | S | P | S | S | S |
| 112989 | C | OPLDDDIH_00099 | hypothetical protein [PA0088] | + | 226 | 1 | CTG | L | L | L | L | L | L | L | V | V | V | V | V | L | V | V | V | L | L | V | L | L | L |
| 114441 | A | OPLDDDIH_00099 | hypothetical protein [PA0088] | + | 1678 | 1 | ACC | T | T | T | T | T | T | T | T | T | A | T | T | T | T | T | T | T | T | T | T | T | T |
| 118185 | C | OPLDDDIH_00101 | ClpV1 [PA0090] | + | 2560 | 1 | CGG | R | R | R | R | R | W | R | R | R | R | R | R | R | R | R | R | R | R | R | R | R | R |
| 121899 | C | OPLDDDIH_00107 | hypothetical protein [PA0093] | - | 1085 | 2 | GGC | G | G | G | G | G | G | G | G | G | G | G | G | G | D | D | G | G | G | G | G | G | G |
| 142314 | A | OPLDDDIH_00126 | hypothetical protein [PA0112] | + | 926 | 2 | AAC | N | N | N | S | N | N | N | N | N | N | N | N | N | N | N | N | N | N | X | N | N | N |
| 147851 | C | OPLDDDIH_00133 | putative dicarboxylate transporter [PA0119] | + | 850 | 1 | CAC | H | H | H | H | H | H | H | H | H | H | H | H | H | H | H | H | H | H | H | H | Y | H |
| 148626 | C | OPLDDDIH_00134 | putative transcriptional regulator [PA0120] | + | 242 | 2 | GCG | A | A | A | A | A | V | A | A | A | A | A | A | A | A | A | A | A | A | A | A | A | A |
| 151310 | G | OPLDDDIH_00137 | putative transcriptional regulator [PA0123] | - | 127 | 1 | CGC | R | R | R | R | R | C | R | R | R | R | R | R | R | R | R | R | R | R | R | R | R | R |
| 172077 | A | OPLDDDIH_00157 | nonspecific ribonucleoside hydrolase [PA0143] | + | 502 | 1 | AAC | N | N | N | N | N | D | N | N | N | N | N | N | N | N | N | N | N | N | N | N | N | N |
| 173165 | C | OPLDDDIH_00158 | Nucleoside-specific channel-forming protein Tsx | - | 349 | 1 | GAA | E | E | E | E | E | E | K | E | E | E | E | E | E | E | E | E | E | E | E | E | E | E |
| 195366 | C | OPLDDDIH_00177 | putative transcriptional regulator [PA0163] | - | 481 | 1 | GAG | E | E | E | E | E | E | E | K | K | K | K | K | E | E | E | E | E | E | E | E | E | E |
| 226356 | C | OPLDDDIH_00204 | putative acid phosphatase [PA0190] | + | 95 | 2 | CCG | P | P | P | L | P | L | L | L | L | L | L | L | P | L | L | L | P | X | X | P | X | P |
| 250462 | C | OPLDDDIH_00228 | putative aldehyde dehydrogenase [PA0219] | + | 722 | 2 | ACC | T | T | T | T | T | T | T | T | T | T | T | T | T | T | T | T | T | T | I | T | T | T |
| 263174 | T | OPLDDDIH_00239 | 3-carboxy-cis,cis-muconate cycloisomerase [PA0230] | + | 869 | 2 | GTG | V | V | V | A | V | V | V | V | V | V | V | V | V | V | V | V | V | V | V | V | V | V |
| 263482 | G | OPLDDDIH_00239 | 3-carboxy-cis,cis-muconate cycloisomerase [PA0230] | + | 1177 | 1 | GAA | E | E | E | E | E | E | E | K | K | K | K | K | E | E | E | E | E | E | E | E | E | E |
| 273340 | C | OPLDDDIH_00249 | putative porin [PA0240] | - | 874 | 1 | GGC | G | G | G | G | G | S | G | G | G | G | G | G | G | G | G | G | G | G | G | G | G | G |
| 279245 | C | OPLDDDIH_00253 | hypothetical protein [PA0244] | - | 466 | 1 | GTC | V | V | V | V | V | V | V | I | I | I | I | I | V | V | V | V | V | V | V | V | V | V |
| 326740 | C | OPLDDDIH_00298 | conserved hypothetical protein [PA0285] | - | 164 | 2 | GGC | G | G | G | D | G | G | G | G | G | G | G | G | G | G | G | G | G | G | G | G | G | G |
| 344162 | G | OPLDDDIH_00312 | putrescine aminotransferase [PA0299] | + | 905 | 2 | GGG | G | G | G | G | G | G | G | G | G | G | G | G | G | G | G | G | G | E | G | G | G | G |
| 356751 | C | OPLDDDIH_00323 | hypothetical protein [PA0309] | - | 398 | 2 | CGG | R | R | R | R | R | R | R | R | R | Q | R | R | R | R | R | R | R | R | R | R | R | R |
| 370159 | C | OPLDDDIH_00338 | putative binding protein component of ABC transporter [PA0323] | - | 493 | 1 | GCG | A | A | A | A | A | T | A | A | A | A | A | A | A | A | A | A | A | A | A | A | A | A |
| 408997 | T | OPLDDDIH_00374 | formamidopyrimidine-DNA glycosylase [PA0357] | + | 788 | 2 | GTG | V | V | V | V | V | V | V | A | A | A | A | A | V | V | V | V | V | V | V | V | V | V |
| 479117 | C | OPLDDDIH_00444 | multidrug resistance operon repressor MexR [PA0424] | - | 301 | 1 | GGG | G | G | W | W | W | W | W | W | W | W | W | W | W | W | W | W | W | W | W | W | W | W |
| 482716 | C | OPLDDDIH_00446 | Resistance-Nodulation-Cell Division (RND) multidrug efflux transporter MexB [PA0426] | + | 1858 | 1 | CGC | R | R | C | C | C | C | C | C | C | C | C | C | C | C | C | C | C | C | C | C | C | C |
| 485808 | C | OPLDDDIH_00448 | putative ATP-dependent RNA helicase [PA0428] | - | 1666 | 1 | GGC | G | G | G | G | G | G | G | G | G | G | G | G | G | G | G | G | G | G | G | G | S | G |
| 492974 | A | OPLDDDIH_00454 | hypothetical protein [PA0434] | + | 344 | 2 | TAC | Y | Y | Y | Y | Y | Y | Y | Y | Y | Y | Y | Y | Y | Y | Y | Y | Y | Y | C | Y | Y | Y |
| 498817 | A | OPLDDDIH_00458 | cytosine permease [PA0438] | - | 749 | 2 | GTG | V | V | V | V | V | A | V | V | V | V | V | V | V | V | V | V | V | V | V | V | V | V |
| 527303 | C | OPLDDDIH_00478 | hypothetical protein [PA0460] | + | 434 | 2 | ACC | T | T | T | T | T | I | T | T | T | T | T | T | T | T | T | T | T | T | T | T | T | T |
| 532083 | C | OPLDDDIH_00483 | inner membrane protein CreD [PA0465] | + | 599 | 2 | GCG | A | A | A | A | A | A | A | A | A | A | A | A | A | A | A | A | A | A | V | A | A | A |
| 534551 | C | OPLDDDIH_00487 | hypothetical protein [PA0468] | - | 379 | 1 | GTA | V | V | V | V | V | V | V | V | V | V | V | V | V | L | L | V | V | V | V | V | V | V |
| 570379 | C | OPLDDDIH_00524 | putative acyl-CoA dehydrogenase [PA0507] | + | 250 | 1 | CAG | Q | Q | Q | Q | Q | Q | Q | Q | Q | Q | Q | Q | Q | Q | Q | Q | Q | Q | Q | Q | * | Q |
| 572429 | C | OPLDDDIH_00525 | putative acyl-CoA dehydrogenase [PA0508] | + | 230 | 2 | CCG | P | P | P | P | P | P | P | P | P | P | P | P | P | L | L | L | P | P | X | P | P | P |
| 607586 | A | OPLDDDIH_00564 | methionine adenosyltransferase [PA0546] | - | 509 | 2 | GTG | V | V | V | V | V | A | V | V | V | V | V | V | V | V | V | V | V | V | V | V | V | V |
| 618854 | A | OPLDDDIH_00576 | conserved hypothetical protein [PA0558] | + | 50 | 2 | CAG | Q | Q | Q | Q | Q | Q | Q | Q | Q | Q | Q | Q | Q | Q | Q | R | Q | Q | Q | Q | Q | Q |
| 626198 | A | OPLDDDIH_00587 | hypothetical protein [PA0570] | - | 47 | 2 | CTG | L | L | L | P | L | L | L | P | P | P | P | P | L | X | X | P | L | X | X | L | P | L |
| 626562 | C | OPLDDDIH_00588 | hypothetical protein [PA0571] | - | 287 | 2 | GGC | G | G | G | G | G | G | G | G | G | G | G | G | G | G | G | G | G | D | G | G | G | G |
| 640596 | A | OPLDDDIH_00599 | conserved hypothetical protein [PA0575] | - | 3308 | 2 | CTG | L | L | L | L | L | L | L | L | L | L | L | L | L | L | L | L | L | P | L | L | L | L |
| 648069 | A | OPLDDDIH_00602 | conserved hypothetical protein [PA0578] | - | 401 | 2 | GTC | V | V | V | V | V | V | V | V | V | V | V | V | V | V | V | V | V | A | V | V | V | V |
| 659431 | A | OPLDDDIH_00614 | bis(5'-nucleosyl)-tetraphosphatase [PA0590] | - | 368 | 2 | ATC | I | I | I | T | I | I | I | T | T | T | T | T | I | T | T | T | I | T | X | I | T | I |
| 692255 | A | OPLDDDIH_00646 | Arsenical-resistance protein Acr3 | - | 958 | 1 | TTC | F | F | F | F | F | F | F | F | F | F | L | F | F | F | F | F | F | F | F | F | F | F |
| 713786 | A | OPLDDDIH_00671 | putative sulfate transporter [PA2563] | + | 545 | 2 | GAC | D | D | D | D | D | D | D | G | G | G | G | G | D | D | D | D | D | D | D | D | D | D |
| 724828 | A | OPLDDDIH_00682 | hypothetical protein | + | 305 | 2 | GAC | D | D | D | D | D | D | D | D | D | D | D | D | D | G | G | D | D | D | D | D | D | D |
| 750320 | C | OPLDDDIH_00714 | putative bacteriophage protein [PA0641] | + | 536 | 2 | CCG | P | P | P | P | P | P | L | P | P | P | P | P | P | P | P | P | P | P | P | P | P | P |
| 759503 | A | OPLDDDIH_00722 | transcriptional regulator Vfr [PA0652] | - | 158 | 2 | ATC | I | I | I | I | I | I | I | T | T | T | T | T | I | T | T | T | I | T | X | I | T | I |
| 761594 | A | OPLDDDIH_00725 | hypothetical protein [PA0655] | - | 577 | 1 | TTC | F | F | F | F | F | F | F | L | L | L | L | L | F | F | F | F | F | F | F | F | F | F |
| 772438 | A | OPLDDDIH_00737 | conserved hypothetical protein [PA0667] | - | 725 | 2 | ATC | I | I | I | I | I | I | I | I | I | I | I | I | I | I | I | I | I | N | I | I | I | I |
| 818631 | T | OPLDDDIH_00791 | excinuclease ABC subunit A [PA4234] | - | 1435 | 1 | ACC | T | T | T | T | T | T | A | T | T | T | T | T | T | T | T | T | T | T | T | X | T | T |
| 866491 | A | OPLDDDIH_00823 | hypothetical protein [PA4202] | + | 826 | 1 | AGC | S | S | S | S | S | S | S | S | S | S | S | S | S | G | G | G | S | S | G | S | S | S |
| 868450 | A | OPLDDDIH_00825 | hypothetical protein [PA4200] | - | 314 | 2 | CTG | L | L | L | L | L | L | L | L | L | L | L | L | L | P | P | L | L | L | L | L | L | L |
| 868537 | A | OPLDDDIH_00825 | hypothetical protein [PA4200] | - | 227 | 2 | GTC | V | V | V | V | V | V | V | V | V | V | V | V | V | V | V | A | V | V | V | V | V | V |
| 929447 | C | OPLDDDIH_00876 | putative toxin transporter [PA4143] | - | 94 | 1 | GCG | A | A | A | A | A | T | A | A | A | A | A | A | A | A | A | A | A | A | A | A | A | A |
| 929938 | A | OPLDDDIH_00877 | putative secretion protein [PA4142] | - | 869 | 2 | GTT | V | V | V | V | V | V | V | V | V | V | V | V | V | V | V | V | V | V | X | V | A | V |
| 953232 | A | OPLDDDIH_00897 | conserved hypothetical protein [PA4122] | - | 211 | 1 | TTC | F | F | F | F | F | F | F | F | F | F | F | F | F | F | F | F | F | F | L | F | F | F |
| 977134 | C | OPLDDDIH_00916 | hypothetical protein [PA4099] | - | 565 | 1 | GGC | G | G | G | G | G | G | G | G | G | G | G | G | G | G | G | G | G | G | G | G | S | G |
| 984451 | A | OPLDDDIH_00924 | 4-hydroxyphenylacetate 3-monooxygenase large chain [PA4091] | - | 901 | 1 | TTC | F | F | F | F | F | F | L | F | F | F | F | F | F | F | F | F | F | F | F | F | F | F |
| 990564 | G | OPLDDDIH_00930 | chaperone CupB2 [PA4085] | + | 499 | 1 | GGG | G | G | G | R | G | G | G | G | G | G | G | G | G | G | G | G | G | G | G | G | G | G |
| 991305 | C | OPLDDDIH_00931 | usher CupB3 [PA4084] | + | 418 | 1 | CGA | R | R | R | R | R | R | R | R | R | R | R | R | R | R | * | R | R | R | R | R | R | R |
| 996462 | A | OPLDDDIH_00933 | adhesive protein CupB5 [PA4082] | + | 1970 | 2 | AAC | N | N | N | N | N | S | N | N | N | N | N | N | N | N | N | N | N | N | N | N | N | N |
| 1002592 | C | OPLDDDIH_00937 | putative nonribosomal peptide synthetase [PA4078] | - | 1261 | 1 | GCG | A | A | A | A | A | A | A | A | A | A | A | A | A | T | X | A | A | A | A | A | A | A |
| 1013492 | A | OPLDDDIH_00948 | hypothetical protein | - | 250 | 1 | TTC | F | F | F | F | F | L | F | F | F | F | F | F | F | F | F | F | F | F | F | F | F | F |
| 1015958 | G | OPLDDDIH_00951 | hypothetical protein [PA4065] | - | 515 | 2 | ACG | T | T | T | M | T | T | T | M | M | M | M | M | T | M | M | M | T | M | M | T | M | T |
| 1024111 | A | OPLDDDIH_00961 | GTP cyclohydrolase II / 3,4-dihydroxy-2-butanone 4-phosphate synthase [PA4054] | + | 782 | 2 | GAC | D | D | D | G | D | D | D | D | D | D | D | D | D | D | D | D | D | D | D | D | D | D |
| 1028147 | C | OPLDDDIH_00967 | hypothetical protein [PA4048] | + | 398 | 2 | GCG | A | A | A | V | A | V | V | V | V | V | V | V | A | V | V | V | A | V | V | A | V | A |
| 1031942 | C | OPLDDDIH_00971 | 1-deoxyxylulose-5-phosphate synthase [PA4044] | - | 421 | 1 | GCC | A | A | A | A | A | A | A | A | A | A | A | A | A | A | A | A | A | A | A | A | T | A |
| 1084921 | T | OPLDDDIH_01019 | penicillin-binding protein 2 [PA4003] | + | 1768 | 1 | TCC | S | S | P | S | P | S | S | S | S | S | S | S | S | S | S | S | S | S | S | S | S | S |
| 1099456 | C | OPLDDDIH_01033 | leucyl-tRNA synthetase [PA3987] | - | 734 | 2 | GGC | G | G | G | G | G | G | G | D | D | D | D | D | G | D | D | D | G | X | D | G | D | G |
| 1129341 | A | OPLDDDIH_01063 | hypothetical protein [PA3958] | - | 383 | 2 | GTC | V | V | V | V | V | V | V | V | V | V | V | V | V | V | V | A | V | V | V | V | V | V |
| 1133596 | C | OPLDDDIH_01068 | conserved hypothetical protein [PA3953] | - | 107 | 2 | GGC | G | G | G | G | G | G | G | G | G | G | G | G | G | G | G | G | G | G | G | G | D | G |
| 1148224 | G | OPLDDDIH_01080 | hypothetical protein [PA3941] | + | 416 | 2 | CGC | R | R | R | R | R | R | R | R | R | R | R | H | R | R | R | R | R | R | R | R | R | R |
| 1156781 | A | OPLDDDIH_01088 | putative choline transporter [PA3933] | - | 1412 | 2 | CTG | L | L | L | L | L | P | L | L | L | L | L | L | L | L | L | L | L | L | L | L | L | L |
| 1163057 | C | OPLDDDIH_01092 | cyanide insensitive terminal oxidase [PA3929] | + | 839 | 2 | CCG | P | P | P | L | P | P | P | L | L | L | L | L | P | L | L | L | P | L | L | P | L | P |
| 1170312 | A | OPLDDDIH_01098 | hypothetical protein [PA3923] | + | 1249 | 1 | ACC | T | T | T | T | T | T | T | T | T | T | T | T | T | T | T | A | T | T | T | T | T | T |
| 1170784 | T | OPLDDDIH_01098 | hypothetical protein [PA3923] | + | 1721 | 2 | GTG | V | V | V | V | V | V | A | V | V | V | V | V | V | V | V | V | V | V | V | V | V | V |
| 1182659 | T | OPLDDDIH_01108 | molybdopterin converting factor, large subunit [PA3916] | + | 257 | 2 | CTG | L | L | L | L | L | L | L | L | L | L | L | L | L | L | L | P | L | L | L | L | L | L |
| 1191326 | A | OPLDDDIH_01115 | Extracelullar DNA degradation protein, EddB [PA3909] | + | 2243 | 2 | GAC | D | D | D | D | D | D | D | D | D | D | G | D | D | D | D | D | D | D | D | D | D | D |
| 1193094 | C | OPLDDDIH_01117 | hypothetical protein [PA3907] | - | 155 | 2 | CGT | R | R | R | R | R | R | R | R | R | H | R | R | R | R | R | R | R | R | R | R | R | R |
| 1241619 | A | OPLDDDIH_01163 | NAD(P)H-dependent anabolic L-arginine dehydrogenase, DauB [PA3862] | - | 68 | 2 | GTC | V | V | V | V | V | V | V | A | V | V | V | V | V | V | V | V | V | V | V | V | V | V |
| 1247107 | A | OPLDDDIH_01167 | putative amino acid-binding protein [PA3858] | + | 596 | 2 | CAG | Q | Q | Q | Q | Q | Q | Q | Q | Q | Q | Q | Q | Q | Q | Q | Q | Q | R | Q | Q | Q | Q |
| 1321949 | A | OPLDDDIH_01246 | hypothetical protein | - | 506 | 2 | GTT | V | V | V | A | V | V | V | V | V | V | V | V | V | V | V | V | V | V | V | V | V | V |
| 1333786 | C | OPLDDDIH_01254 | ATP-dependent DNA helicase Rep | + | 679 | 1 | CCT | P | P | P | P | P | P | P | P | P | P | P | P | P | P | P | P | P | P | P | P | S | P |
| 1341635 | T | OPLDDDIH_01261 | secretion protein SecD [PA3821] | + | 1123 | 1 | TAC | Y | Y | Y | Y | Y | Y | Y | Y | Y | Y | Y | Y | Y | Y | Y | Y | Y | Y | Y | Y | H | Y |
| 1361021 | A | OPLDDDIH_01283 | conserved hypothetical protein [PA3799] | + | 740 | 2 | TAC | Y | Y | Y | Y | Y | Y | C | Y | Y | Y | Y | Y | Y | Y | Y | Y | Y | Y | Y | Y | Y | Y |
| 1362325 | C | OPLDDDIH_01284 | putative aminotransferase [PA3798] | + | 266 | 2 | ACC | T | T | T | T | T | T | T | T | T | T | T | T | T | T | T | T | T | I | T | T | T | T |
| 1365177 | C | OPLDDDIH_01287 | putative oxidoreductase [PA3795] | - | 574 | 1 | GAC | D | D | D | D | D | D | D | D | D | D | D | D | D | D | D | D | D | N | D | D | D | D |
| 1376135 | A | OPLDDDIH_01298 | hypothetical protein [PA3784] | + | 340 | 1 | ACG | T | T | T | T | T | T | T | T | T | T | T | T | T | T | T | T | T | T | T | T | A | T |
| 1380749 | C | OPLDDDIH_01304 | hypothetical protein [PA3779] | - | 1005 | 3 | ATG | M | M | M | M | M | M | M | M | M | M | I | M | M | M | M | M | M | M | M | M | M | M |
| 1384879 | T | OPLDDDIH_01307 | putative transcriptional regulator [PA3776] | - | 293 | 2 | CAC | H | H | H | R | H | R | R | R | R | R | R | R | H | R | R | R | H | X | X | H | X | H |
| 1386225 | C | OPLDDDIH_01309 | putative acetylpolyamine aminohydrolase [PA3774] | - | 964 | 1 | GGC | G | G | G | G | G | G | S | G | G | G | G | G | G | G | G | G | G | G | X | G | G | G |
| 1387996 | A | OPLDDDIH_01310 | hypothetical protein [PA3773] | - | 380 | 2 | CTC | L | L | L | L | L | L | L | L | L | L | L | L | L | L | L | L | L | P | L | L | L | L |
| 1415165 | A | OPLDDDIH_01332 | phosphoribosylglycinamide formyltransferase 2 [PA3751] | + | 224 | 2 | AAG | K | K | K | K | K | K | K | K | K | K | K | K | K | R | R | R | K | K | R | K | K | K |
| 1415453 | C | OPLDDDIH_01332 | phosphoribosylglycinamide formyltransferase 2 [PA3751] | + | 512 | 2 | CCC | P | P | P | P | P | P | P | P | P | P | P | P | P | P | P | P | P | L | P | P | P | P |
| 1431435 | C | OPLDDDIH_01348 | threonine synthase [PA3735] | + | 178 | 1 | CGC | R | R | R | R | R | R | R | R | R | R | R | R | R | R | R | R | R | R | R | R | C | R |
| 1444157 | C | OPLDDDIH_01356 | hypothetical protein [PA3728] | + | 3836 | 2 | ACG | T | T | T | M | T | T | T | T | T | T | T | T | T | T | T | T | T | T | T | T | T | T |
| 1452128 | C | OPLDDDIH_01362 | hypothetical protein [PA3722] | + | 206 | 2 | GCC | A | A | A | G | A | G | G | G | G | G | G | G | A | G | G | G | A | G | G | A | G | A |
| 1461928 | T | OPLDDDIH_01371 | hypothetical protein [PA3712] | + | 317 | 2 | CTG | L | L | L | P | L | P | P | P | P | P | P | P | L | P | P | P | L | P | X | L | P | L |
| 1473389 | G | OPLDDDIH_01379 | putative chemotaxis sensor/effector fusion protein [PA3704] | + | 2209 | 1 | GGT | G | G | G | S | G | G | G | S | S | S | S | S | G | S | S | S | G | S | S | G | S | G |
| 1530713 | A | OPLDDDIH_01434 | conserved hypothetical protein [PA3649] | + | 166 | 1 | ACC | T | T | T | T | T | T | T | A | T | T | T | T | T | T | T | T | T | T | T | T | T | T |
| 1533597 | A | OPLDDDIH_01435 | outer membrane protein Opr86 [PA3648] | + | 1627 | 1 | ACC | T | T | T | T | T | T | T | T | T | T | A | T | T | T | T | T | T | T | T | T | T | T |
| 1553523 | A | OPLDDDIH_01453 | putative transcriptional regulator [PA3630] | - | 758 | 2 | CTG | L | L | L | L | L | L | P | L | L | L | L | L | L | L | L | L | L | L | L | L | L | L |
| 1558990 | C | OPLDDDIH_01459 | L-isoaspartate protein carboxylmethyltransferase type II [PA3624] | + | 217 | 1 | CCG | P | P | P | P | P | P | P | S | S | S | S | S | P | P | P | P | P | P | P | P | P | P |
| 1565160 | T | OPLDDDIH_01464 | hypothetical protein [PA3619] | - | 848 | 2 | AAC | N | N | N | N | N | N | N | N | N | N | S | N | N | N | N | N | N | N | N | N | N | N |
| 1578877 | A | OPLDDDIH_01477 | conserved hypothetical protein [PA3606] | + | 299 | 2 | GAC | D | D | D | G | D | D | D | D | D | D | D | D | D | D | D | D | D | D | D | D | D | D |
| 1589981 | A | OPLDDDIH_01489 | putative transcriptional regulator [PA3594] | - | 766 | 1 | TAC | Y | Y | Y | Y | Y | H | Y | Y | Y | Y | Y | Y | Y | Y | Y | Y | Y | Y | Y | Y | Y | Y |
| 1594012 | A | OPLDDDIH_01492 | putative enoyl-CoA hydratase/isomerase [PA3591] | + | 140 | 2 | CAC | H | H | H | H | H | H | H | H | H | H | R | H | H | H | H | H | H | H | H | H | H | H |
| 1601916 | A | OPLDDDIH_01499 | glycerol-3-phosphate dehydrogenase [PA3584] | - | 1043 | 2 | CTC | L | L | L | L | L | P | L | L | L | L | L | L | L | L | L | L | L | L | L | L | L | L |
| 1696924 | C | OPLDDDIH_01580 | putative decarboxylase [PA3506] | + | 290 | 2 | ACC | T | T | T | T | T | T | T | T | T | T | T | I | T | T | T | T | T | T | T | T | T | T |

| | | | | | | | | | | | | | | | | | | | | | | | | | | | | | |
|---|---|---|---|---|---|---|---|---|---|---|---|---|---|---|---|---|---|---|---|---|---|---|---|---|---|---|---|---|---|
| 1702712 | A | OPLDDDIH_01586 | conserved hypothetical protein [PA3500] | + | 539 | 2 | CAC | H | H | H | H | H | H | H | H | H | H | H | R | H | H | H | H | H | H | H | H | H | H |
| 1709837 | C | OPLDDDIH_01595 | putative ferredoxin [PA3491] | - | 2147 | 2 | CGG | R | R | R | R | R | Q | R | R | R | R | R | R | R | R | R | R | R | R | X | X | R | R |
| 1742987 | C | OPLDDDIH_01621 | conserved hypothetical protein [PA3465] | - | 1162 | 1 | GGC | G | G | G | G | G | G | G | S | S | S | S | S | G | G | G | G | G | G | G | G | G | G |
| 1743598 | C | OPLDDDIH_01621 | conserved hypothetical protein [PA3465] | - | 551 | 2 | GGC | G | G | G | G | G | G | D | G | G | G | G | G | G | G | G | G | G | G | G | G | G | G |
| 1762893 | A | OPLDDDIH_01634 | malate:quinone oxidoreductase [PA3452] | - | 86 | 2 | GTG | V | V | V | A | V | V | V | V | V | V | V | V | V | V | V | V | V | V | V | V | V | V |
| 1766699 | T | OPLDDDIH_01639 | putative ATP-binding component of ABC transporter [PA3447] | + | 386 | 2 | CTG | L | L | L | L | L | L | L | L | L | L | L | P | L | L | L | L | L | L | L | L | L | L |
| 1778801 | A | OPLDDDIH_01656 | putative aldolase [PA3430] | + | 305 | 2 | CAC | H | H | H | H | H | H | H | H | H | H | H | H | H | R | H | H | H | H | H | H | H | H |
| 1820120 | C | OPLDDDIH_01696 | nitrous-oxide reductase precursor [PA3392] | - | 932 | 2 | AGC | S | S | S | S | S | S | S | S | S | S | S | S | S | S | S | S | S | S | S | S | N | S |
| 1822365 | A | OPLDDDIH_01697 | regulatory protein NosR [PA3391] | - | 877 | 1 | TAT | Y | Y | Y | Y | Y | Y | Y | Y | Y | Y | H | Y | Y | Y | Y | Y | Y | Y | Y | Y | Y | Y |
| 1856554 | A | OPLDDDIH_01736 | D-serine dehydratase [PA3357] | - | 736 | 1 | TTC | F | F | F | L | F | L | L | L | L | L | L | L | F | L | L | L | F | X | X | F | L | F |
| 1857205 | A | OPLDDDIH_01736 | D-serine dehydratase [PA3357] | - | 85 | 1 | TTC | F | F | F | L | F | L | L | L | L | L | L | L | F | L | L | L | F | L | L | F | L | F |
| 1862954 | C | OPLDDDIH_01743 | hypothetical protein [PA3350] | - | 598 | 1 | GCC | A | A | A | A | A | A | A | T | T | T | T | T | A | A | A | A | A | A | A | A | A | A |
| 1873970 | C | OPLDDDIH_01753 | hypothetical protein [PA3340] | + | 265 | 1 | CGC | R | R | R | R | R | R | R | R | R | R | R | R | R | R | R | R | R | C | R | R | R | R |
| 1893599 | A | OPLDDDIH_01766 | putative non-ribosomal peptide synthetase [PA3327] | - | 1321 | 1 | TTC | F | F | F | F | F | F | L | F | F | F | F | F | F | F | F | F | F | F | F | F | F | F |
| 1925959 | A | OPLDDDIH_01795 | hypothetical protein [PA3301] | - | 95 | 2 | CTG | L | L | L | L | L | L | L | L | L | L | L | L | L | L | L | L | L | L | L | L | P | L |
| 1933515 | C | OPLDDDIH_01799 | putative ATP-dependent helicase [PA3297] | + | 3097 | 1 | CCC | P | P | P | P | P | P | P | P | P | P | P | S | P | P | P | P | P | P | P | P | P | P |
| 1948457 | A | OPLDDDIH_01811 | hypothetical protein [PA3284] | + | 116 | 2 | TAC | Y | Y | Y | Y | Y | C | Y | Y | Y | Y | Y | Y | Y | Y | Y | Y | Y | Y | Y | Y | Y | Y |
| 2000533 | C | OPLDDDIH_01862 | hypothetical protein [PA3237] | - | 55 | 1 | GCC | A | A | A | A | A | A | A | T | T | T | T | T | A | A | A | A | A | A | A | A | A | A |
| 2028743 | C | OPLDDDIH_01894 | putative permease of ABC transporter [PA3211] | - | 1108 | 1 | GCC | A | A | A | A | A | A | A | T | T | T | T | T | A | A | A | A | A | A | A | A | A | A |
| 2032095 | C | OPLDDDIH_01897 | conserved hypothetical protein [PA3208] | + | 131 | 2 | CCC | P | P | P | P | P | L | P | P | P | P | P | P | P | P | P | P | P | P | P | P | P | P |
| 2036639 | C | OPLDDDIH_01904 | conserved hypothetical protein [PA3201] | - | 529 | 1 | GGC | G | G | G | G | G | G | S | G | G | G | G | G | G | G | G | G | G | G | G | G | G | G |
| 2041487 | C | OPLDDDIH_01909 | hypothetical protein [PA3196] | + | 448 | 1 | CCG | P | P | P | P | P | P | P | P | P | P | P | P | P | S | S | P | P | P | P | P | P | P |
| 2059652 | C | OPLDDDIH_01926 | hypothetical protein [PA3180] | - | 245 | 2 | GGC | G | G | G | G | G | G | G | G | G | G | G | D | G | G | G | G | G | G | G | G | G | G |
| 2071356 | C | OPLDDDIH_01938 | DNA gyrase subunit A [PA3168] | + | 248 | 2 | ACC | T | T | T | T | T | T | T | T | T | T | T | T | T | I | I | I | T | I | I | T | I | T |
| 2085711 | A | OPLDDDIH_01948 | hypothetical protein | + | 341 | 2 | GAA | E | E | E | E | E | E | E | E | E | E | E | G | E | E | E | E | E | E | E | E | E | E |
| 2093920 | A | OPLDDDIH_01955 | glycosyltransferase WbpL [PA3145] | + | 757 | 1 | ACC | T | T | A | T | T | T | T | T | T | T | T | T | T | T | T | T | T | T | T | T | T | T |
| 2102451 | C | OPLDDDIH_01962 | putative secretion protein [PA3136] | - | 469 | 1 | GCC | A | A | A | A | A | A | T | A | A | A | A | A | A | A | A | A | A | A | A | A | A | A |
| 2104929 | A | OPLDDDIH_01964 | glutamyl-tRNA synthetase [PA3134] | + | 950 | 2 | AAG | K | K | K | K | K | K | K | K | K | K | K | K | K | K | K | R | K | K | K | K | K | K |
| 2105385 | A | OPLDDDIH_01964 | glutamyl-tRNA synthetase [PA3134] | + | 1406 | 2 | CAG | Q | Q | Q | Q | Q | Q | Q | Q | Q | Q | Q | Q | Q | Q | Q | Q | Q | R | Q | Q | Q | Q |
| 2115203 | A | OPLDDDIH_01980 | putative transcriptional regulator [PA3122] | - | 800 | 2 | ATC | I | I | I | T | I | I | I | I | I | I | I | I | I | I | I | I | I | I | I | I | I | I |
| 2117638 | C | OPLDDDIH_01982 | 3-isopropylmalate dehydratase small subunit [PA3120] | + | 38 | 2 | CCG | P | P | P | L | P | P | P | L | L | L | L | L | P | L | L | L | P | L | X | P | L | P |
| 2125797 | C | OPLDDDIH_01988 | tRNA-pseudouridine synthase I [PA3114] | + | 341 | 2 | GCG | A | A | A | V | A | A | A | V | V | V | V | V | A | V | X | V | A | V | X | A | V | A |
| 2138981 | G | OPLDDDIH_02000 | general secretion pathway protein F [PA3102] | + | 22 | 1 | GCC | A | A | A | A | A | A | A | A | A | A | A | A | A | T | A | A | A | A | A | A | A | A |
| 2163255 | A | OPLDDDIH_02024 | conserved hypothetical protein [PA3081] | + | 292 | 1 | ACC | T | T | T | T | T | T | T | T | T | T | T | T | T | A | T | T | T | T | T | T | T | T |
| 2172364 | A | OPLDDDIH_02029 | putative serine protease [PA3535] | + | 2005 | 1 | AGC | S | S | S | S | S | S | S | S | S | S | S | S | S | S | S | S | S | S | S | S | G | S |
| 2174621 | T | OPLDDDIH_02031 | hypothetical protein [PA3075] | - | 1435 | 1 | ACC | T | T | T | T | T | T | T | T | T | T | A | T | T | T | T | T | T | T | T | T | T | T |
| 2175562 | A | OPLDDDIH_02031 | hypothetical protein [PA3075] | - | 494 | 2 | CTG | L | L | L | L | L | L | L | L | L | L | L | L | L | L | L | L | L | L | L | L | P | L |
| 2179590 | C | OPLDDDIH_02035 | hypothetical protein [PA3071] | - | 664 | 1 | GAC | D | D | D | D | D | D | D | N | N | N | N | N | D | N | N | N | D | N | X | D | N | D |
| 2203063 | C | OPLDDDIH_02051 | hypothetical protein [PA3056] | + | 295 | 1 | CCG | P | P | P | P | P | P | P | P | P | P | P | P | P | P | P | S | P | P | P | P | P | P |
| 2214434 | C | OPLDDDIH_02061 | conserved hypothetical protein [PA3046] | + | 325 | 1 | CCT | P | P | P | P | P | S | P | P | P | P | P | P | P | P | P | P | P | P | P | P | P | P |
| 2216913 | C | OPLDDDIH_02063 | putative two-component sensor [PA3044] | + | 1396 | 1 | CGG | R | R | R | W | R | W | W | W | W | W | W | W | R | W | W | W | R | W | W | R | W | R |
| 2274244 | A | OPLDDDIH_02117 | soluble pyridine nucleotide transhydrogenase [PA2991] | + | 427 | 1 | ATC | I | I | I | V | I | V | V | V | V | V | V | V | I | V | V | V | I | V | V | I | V | I |
| 2274275 | C | OPLDDDIH_02117 | soluble pyridine nucleotide transhydrogenase [PA2991] | + | 458 | 2 | TCG | S | S | S | S | S | L | S | S | S | S | S | S | S | S | S | S | S | S | S | S | S | S |
| 2276044 | C | OPLDDDIH_02119 | hypothetical protein | - | 239 | 2 | CGG | R | R | R | R | R | R | R | R | R | R | R | R | R | R | Q | R | R | R | R | R | R | R |
| 2287496 | C | OPLDDDIH_02132 | UDP-N-acetylpyruvoylglucosamine reductase [PA2977] | + | 695 | 2 | GCC | A | A | A | A | A | A | A | A | A | A | A | A | A | A | V | A | A | A | A | A | A | A |
| 2287682 | T | OPLDDDIH_02132 | UDP-N-acetylpyruvoylglucosamine reductase [PA2977] | + | 881 | 2 | GTA | V | V | V | V | V | V | V | V | V | V | V | V | G | V | V | V | V | V | V | V | V | V |
| 2299612 | C | OPLDDDIH_02144 | beta-ketoacyl-acyl carrier protein synthase II [PA2965] | + | 380 | 2 | CCG | P | P | P | P | P | P | P | P | P | P | P | P | P | L | L | P | P | P | P | P | P | P |
| 2320509 | C | OPLDDDIH_02166 | cobalamin biosynthetic protein CobN [PA2944] | + | 818 | 2 | CCG | P | P | P | P | P | P | P | P | P | L | P | P | P | P | P | P | P | P | P | P | P | P |
| 2334891 | A | OPLDDDIH_02179 | putative major facilitator superfamily (MFS) transporter [PA2933] | - | 646 | 1 | TTC | F | F | F | F | F | F | F | F | F | F | F | F | F | F | F | F | F | F | F | F | L | F |
| 2339548 | T | OPLDDDIH_02185 | hypothetical protein | + | 380 | 2 | GTA | V | V | V | V | V | V | V | V | V | V | A | V | V | V | V | V | V | V | V | V | V | V |
| 2360332 | C | OPLDDDIH_02205 | cobalamin biosynthetic protein CbiD [PA2908] | - | 955 | 1 | GCC | A | A | A | T | A | A | A | A | A | A | A | A | A | A | A | A | A | A | A | A | A | A |
| 2366748 | A | OPLDDDIH_02210 | precorrin-3 methylase CobJ [PA2903] | + | 1061 | 2 | CAG | Q | Q | Q | Q | Q | Q | Q | Q | Q | Q | R | Q | Q | Q | Q | Q | Q | Q | Q | Q | Q | Q |
| 2371295 | G | OPLDDDIH_02216 | putative transcriptional regulator [PA2897] | + | 337 | 1 | GGC | G | G | G | G | G | G | G | G | G | G | G | G | G | G | G | G | G | G | G | G | S | G |
| 2372966 | A | OPLDDDIH_02217 | putative sigma-70 factor, ECF subfamily [PA2896] | + | 289 | 1 | ACC | T | T | T | A | T | T | T | T | T | T | T | T | T | T | T | T | T | T | T | T | T | T |
| 2373776 | C | OPLDDDIH_02218 | hypothetical protein [PA2895] | + | 467 | 2 | GCG | A | A | A | V | A | A | A | V | V | V | V | V | A | V | X | V | A | X | X | A | X | A |
| 2392066 | C | OPLDDDIH_02234 | putative transcriptional regulator [PA2879] | + | 788 | 2 | CCG | P | P | P | P | P | L | P | P | P | P | P | P | P | P | P | P | P | X | P | P | P | P |
| 2394435 | A | OPLDDDIH_02237 | orotidine 5'-phosphate decarboxylase [PA2876] | + | 566 | 2 | GAC | D | D | D | D | D | D | D | D | D | D | D | D | D | D | D | D | D | D | D | D | G | D |
| 2421972 | A | OPLDDDIH_02268 | conserved hypothetical protein [PA2847] | + | 139 | 1 | ACC | T | T | T | T | T | T | T | T | T | T | T | T | T | T | T | A | T | T | T | T | T | T |
| 2432618 | C | OPLDDDIH_02278 | putative outer membrane protein precursor [PA2837] | - | 1262 | 2 | CGC | R | R | R | R | R | R | R | R | R | R | R | H | R | R | R | R | R | R | R | R | R | R |
| 2446008 | C | OPLDDDIH_02291 | putative sensor/response regulator hybrid [PA2824] | - | 992 | 2 | AGC | S | S | S | N | S | N | N | N | N | N | N | N | S | X | X | N | S | X | X | S | N | S |
| 2447488 | A | OPLDDDIH_02292 | conserved hypothetical protein [PA2823] | + | 407 | 2 | CAA | Q | Q | Q | Q | Q | Q | Q | Q | Q | Q | Q | R | Q | Q | Q | Q | Q | Q | Q | Q | Q | Q |
| 2451453 | C | OPLDDDIH_02301 | hypothetical protein | - | 343 | 1 | GGC | G | G | G | G | G | G | G | G | G | G | G | G | G | G | G | G | G | G | G | G | S | G |
| 2462822 | A | OPLDDDIH_02314 | putative ATP-binding component of ABC transporter [PA2812] | + | 632 | 2 | CAC | H | H | H | H | H | H | H | H | H | H | H | H | H | H | H | H | H | H | H | H | R | H |
| 2468173 | C | OPLDDDIH_02321 | hypothetical protein [PA2805] | - | 169 | 1 | GTC | V | V | V | V | V | V | I | V | V | V | V | V | V | V | V | V | V | V | V | V | V | V |
| 2473423 | A | OPLDDDIH_02328 | putative two-component response regulator [PA2798] | + | 1019 | 2 | GAT | D | D | D | D | D | D | D | D | D | D | D | D | D | D | D | D | D | D | G | D | D | D |
| 2517683 | A | OPLDDDIH_02374 | conserved hypothetical protein [PA2776] | - | 1202 | 2 | TTC | F | F | F | F | F | F | F | F | F | F | F | F | F | S | F | F | F | F | F | F | F | F |
| 2525525 | C | OPLDDDIH_02386 | putative enoyl-CoA hydratase/isomerase [PA2767] | - | 532 | 1 | GGC | G | G | G | G | G | G | G | S | G | G | G | G | G | G | G | G | G | G | G | G | G | G |
| 2587546 | G | OPLDDDIH_02460 | hypothetical protein | + | 106 | 1 | GCC | A | A | A | A | A | A | A | A | A | A | A | A | A | A | A | A | A | A | T | A | A | A |
| 2632331 | A | OPLDDDIH_02496 | hypothetical protein | + | 1301 | 2 | AAC | N | N | N | N | N | N | N | N | N | N | N | N | N | N | N | N | N | N | N | N | S | N |
| 2642558 | C | OPLDDDIH_02503 | hypothetical protein | - | 2981 | 2 | CGC | R | R | R | R | R | H | R | R | R | R | R | R | R | R | R | R | R | R | R | R | R | R |
| 2729348 | A | OPLDDDIH_02584 | hypothetical protein [PA5471] | - | 514 | 1 | TGG | W | W | W | W | W | R | W | W | W | W | W | W | W | W | W | W | W | W | W | W | W | W |
| 2765556 | C | OPLDDDIH_02615 | ATP-binding protease component ClpA [PA2620] | + | 1505 | 2 | ACC | T | T | T | T | T | T | T | T | T | T | T | T | T | T | T | T | T | T | T | T | I | T |
| 2766063 | T | OPLDDDIH_02615 | ATP-binding protease component ClpA [PA2620] | + | 2012 | 2 | CTC | L | L | L | L | L | L | L | P | P | L | P | P | L | L | L | L | L | L | L | L | L | L |
| 2782745 | C | OPLDDDIH_02633 | putative thiosulfate sulfurtransferase [PA2603] | - | 20 | 2 | CGC | R | R | R | R | R | R | R | R | R | R | R | R | R | R | R | R | R | R | R | R | H | R |
| 2793040 | C | OPLDDDIH_02644 | putative periplasmic spermidine/putrescine-binding protein [PA2592] | - | 1054 | 1 | GTC | V | V | V | I | V | V | V | I | I | I | I | I | V | I | I | I | V | X | I | V | I | V |
| 2880575 | A | OPLDDDIH_02718 | putative Resistance-Nodulation-Cell Division (RND) efflux transporter [PA2527] | + | 3016 | 1 | ACC | T | T | T | T | T | T | T | T | T | T | T | T | T | T | T | T | T | A | T | T | T | T |
| 2893026 | C | OPLDDDIH_02726 | Resistance-Nodulation-Cell Division (RND) divalent metal cation efflux transporter CzcA [PA2520] | + | 1940 | 2 | GCG | A | A | A | A | A | A | A | A | A | A | A | A | A | A | A | A | A | A | A | A | V | A |
| 2931446 | C | OPLDDDIH_02765 | hypothetical protein [PA2481] | + | 659 | 2 | TCC | S | S | S | F | S | S | S | F | F | F | F | F | S | F | F | F | S | F | F | S | F | S |
| 2942729 | A | OPLDDDIH_02776 | gentisate 1,2-dioxygenase [PA2470] | - | 854 | 2 | GTC | V | V | V | V | V | V | V | V | V | V | V | A | V | V | V | V | V | V | V | V | V | V |
| 3015620 | A | OPLDDDIH_02817 | sigma factor PvdS [PA2426] | - | 212 | 2 | GTC | V | V | A | A | A | A | A | A | A | A | A | A | A | A | A | A | A | A | A | A | A | A |
| 3016662 | C | OPLDDDIH_02818 | PvdG [PA2425] | + | 188 | 2 | GCC | A | A | A | A | A | A | A | V | V | V | V | V | A | A | A | A | A | A | A | A | A | A |
| 3032839 | A | OPLDDDIH_02822 | hypothetical protein [PA2421] | - | 677 | 2 | ATC | I | I | I | T | I | I | I | T | T | T | T | T | I | T | T | T | I | T | T | I | T | I |
| 3042368 | A | OPLDDDIH_02830 | L-2,4-diaminobutyrate:2-ketoglutarate 4-aminotransferase, PvdH [PA2413] | + | 728 | 2 | GAG | E | E | E | E | E | E | E | E | E | E | E | E | E | G | G | E | E | E | E | E | E | E |
| 3099165 | C | OPLDDDIH_02857 | 3-oxo-C12-homoserine lactone acylase PvdQ [PA2385] | + | 1171 | 1 | CGC | R | R | R | R | R | R | R | R | R | R | R | C | R | R | R | R | R | R | R | R | R | R |
| 3186065 | A | OPLDDDIH_02931 | AmbB [PA2305] | + | 2783 | 2 | CAC | H | H | H | H | H | R | H | H | H | H | H | X | H | H | H | H | H | X | X | H | H | H |
| 3187967 | C | OPLDDDIH_02932 | AmbC [PA2304] | + | 907 | 1 | CGG | R | R | R | R | R | W | R | R | R | R | R | R | R | R | R | R | R | R | R | R | R | R |
| 3195489 | A | OPLDDDIH_02934 | AmbE [PA2302] | + | 6305 | 2 | GAG | E | E | E | E | E | E | E | G | E | E | E | E | E | E | E | E | E | X | X | X | E | E |
| 3202098 | T | OPLDDDIH_02941 | hypothetical protein [PA2293] | + | 956 | 2 | GTG | V | V | V | V | V | V | V | V | V | V | V | V | V | V | V | V | V | V | A | V | V | V |
| 3213238 | A | OPLDDDIH_02951 | hypothetical protein [PA2283] | - | 1049 | 2 | TTC | F | F | F | F | F | F | F | F | F | F | F | F | F | F | F | F | F | F | F | F | S | F |
| 3231319 | C | OPLDDDIH_02969 | gluconate dehydrogenase [PA2265] | - | 115 | 1 | GGC | G | G | G | S | G | S | S | S | S | S | S | S | G | S | S | S | G | X | S | G | S | G |
| 3245809 | A | OPLDDDIH_02982 | putative AGCS sodium/alanine/glycine symporter [PA2252] | - | 130 | 1 | TTC | F | F | F | F | F | F | L | F | F | F | F | F | F | F | F | F | F | F | F | F | F | F |
| 3250105 | C | OPLDDDIH_02986 | 2-oxoisovalerate dehydrogenase (beta subunit) [PA2248] | - | 691 | 1 | GGC | G | G | G | G | G | G | G | G | G | G | G | G | G | G | G | G | G | G | G | S | G | G |
| 3250582 | A | OPLDDDIH_02986 | 2-oxoisovalerate dehydrogenase (beta subunit) [PA2248] | - | 214 | 1 | TCC | S | S | S | S | S | S | S | S | S | S | S | S | S | S | S | S | S | S | P | S | S | S |
| 3322719 | T | OPLDDDIH_03051 | transport protein ExbD [PA0199] | + | 320 | 2 | GTC | V | V | V | V | V | V | V | V | V | V | V | V | G | V | V | V | V | V | V | V | V | V |
| 3343496 | C | OPLDDDIH_03064 | hypothetical protein | - | 656 | 2 | CGG | R | R | R | R | R | R | R | R | R | R | R | R | R | R | R | Q | R | R | R | R | R | R |
| 3369392 | C | OPLDDDIH_03085 | Resistance-Nodulation-Cell Division (RND) divalent metal cation efflux transporter CzcA [PA2520] | + | 3112 | 1 | CAG | Q | Q | Q | Q | Q | Q | Q | Q | Q | Q | Q | Q | Q | Q | Q | Q | Q | * | Q | Q | Q | Q |
| 3372441 | T | OPLDDDIH_03090 | hypothetical protein | - | 425 | 2 | TAC | Y | Y | Y | C | Y | Y | Y | C | C | C | C | C | Y | C | C | C | Y | C | X | Y | C | Y |
| 3416123 | A | OPLDDDIH_03129 | betaine aldehyde dehydrogenase [PA5373] | + | 529 | 1 | ACG | T | T | T | A | T | T | T | T | T | T | T | T | T | T | T | T | T | T | T | T | T | T |
| 3416472 | A | OPLDDDIH_03129 | betaine aldehyde dehydrogenase [PA5373] | + | 878 | 2 | TAC | Y | Y | Y | Y | Y | Y | Y | Y | Y | Y | Y | Y | Y | Y | Y | Y | Y | Y | Y | Y | C | Y |
| 3444268 | C | OPLDDDIH_03152 | hypothetical protein [PA2209] | + | 691 | 1 | CAC | H | H | H | H | H | H | H | Y | Y | Y | Y | X | H | X | X | Y | H | H | X | H | H | H |
| 3458419 | C | OPLDDDIH_03167 | hydrogen cyanide synthase HcnB [PA2194] | - | 896 | 2 | AGC | S | S | S | S | S | N | S | S | S | S | S | S | S | S | S | S | S | S | S | S | S | S |
| 3463269 | T | OPLDDDIH_03172 | hypothetical protein [PA2181] | - | 356 | 2 | GAG | E | E | E | E | E | E | E | E | E | G | E | E | E | E | E | E | E | E | X | E | E | E |
| 3520724 | C | OPLDDDIH_03228 | usher CupA3 [PA2130] | - | 2075 | 2 | GGC | G | G | G | G | G | G | G | G | G | G | G | G | G | G | G | G | G | G | G | G | A | G |
| 3566011 | C | OPLDDDIH_03268 | putative asparagine synthetase [PA2084] | - | 493 | 1 | GCG | A | A | A | A | A | A | A | A | A | A | A | A | A | A | A | A | A | A | A | A | T | A |
| 3578287 | C | OPLDDDIH_03278 | hypothetical protein [PA2075] | + | 212 | 2 | GCC | A | A | A | V | A | A | A | V | V | V | V | V | A | X | X | V | A | X | X | A | V | A |
| 3579156 | A | OPLDDDIH_03278 | hypothetical protein [PA2075] | + | 1081 | 1 | ACC | T | T | T | T | T | A | T | T | T | T | T | T | T | T | T | T | T | T | T | T | T | T |
| 3594898 | T | OPLDDDIH_03287 | copper resistance protein B precursor [PA2064] | + | 541 | 1 | TGG | W | W | W | R | W | W | W | W | W | W | W | W | W | W | W | W | W | W | W | W | W | W |
| 3624250 | G | OPLDDDIH_03310 | putative amino acid permease [PA2041] | - | 1046 | 2 | GCG | A | A | A | A | A | A | A | A | A | A | A | A | A | A | A | A | A | A | X | A | V | A |
| 3632182 | T | OPLDDDIH_03316 | hypothetical protein [PA2034] | - | 40 | 1 | ACG | T | T | T | T | T | T | T | T | T | T | T | T | T | T | T | T | T | A | T | T | T | T |
| 3642947 | G | OPLDDDIH_03330 | putative transcriptional regulator [PA2020] | - | 400 | 1 | CAA | Q | Q | Q | Q | Q | Q | * | Q | Q | Q | Q | Q | Q | Q | Q | Q | Q | Q | Q | Q | Q | Q |
| 3665431 | G | OPLDDDIH_03348 | 3-hydroxybutyrate dehydrogenase [PA2003] | + | 265 | 1 | GGC | G | G | G | S | G | S | S | S | S | S | S | S | G | S | S | S | G | S | X | G | S | G |
| 3692281 | G | OPLDDDIH_03375 | hypothetical protein [PA1977] | + | 139 | 1 | GCC | A | A | A | T | A | A | A | A | A | A | A | A | A | A | A | A | A | A | A | A | A | A |
| 3709736 | T | OPLDDDIH_03388 | hypothetical protein | - | 13 | 1 | ACT | T | T | T | T | T | T | T | T | T | T | T | T | T | T | T | T | T | A | T | T | T | T |
| 3717796 | T | OPLDDDIH_03394 | hypothetical protein | - | 2192 | 2 | TAC | Y | Y | Y | Y | Y | Y | Y | C | C | C | C | C | Y | Y | Y | Y | Y | Y | Y | Y | Y | Y |
| 3725195 | T | OPLDDDIH_03402 | hypothetical protein | - | 223 | 1 | ACC | T | T | T | T | T | T | T | T | T | T | T | T | T | A | A | A | T | T | A | T | T | T |
| 3734725 | T | OPLDDDIH_03415 | Putative DNA repair helicase RadD | - | 112 | 1 | ACG | T | T | T | T | T | T | T | A | T | T | T | T | T | T | T | T | T | T | T | T | T | T |

| | | | | | | | | | | | | | | | | | | | | | | | | | | | | |
|---|---|---|---|---|---|---|---|---|---|---|---|---|---|---|---|---|---|---|---|---|---|---|---|---|---|---|---|---|
| 3737374 G | OPLDDDIH_03421 | hypothetical protein | + | 97 | 1 | GGC | G | G | G | G | G | G | G | G | G | G | G | G | G | G | G | S | G | G | G | G | G | G |
| 3777453 A | OPLDDDIH_03467 | hypothetical protein [PA1941] | + | 1693 | 1 | AGC | S | S | S | S | S | S | S | S | S | S | S | S | S | S | S | G | S | S | S | S | S | S |
| 3791562 T | OPLDDDIH_03479 | conserved hypothetical protein [PA1926] | - | 2048 | 2 | GAC | D | D | D | D | D | D | D | G | G | G | G | G | D | D | D | D | D | D | D | D | D | D |
| 3798741 G | OPLDDDIH_03483 | putative TonB-dependent receptor [PA1922] | - | 1552 | 1 | CGG | R | R | R | R | R | W | R | R | R | R | R | R | R | R | R | R | R | R | R | R | R | R |
| 3800180 G | OPLDDDIH_03483 | putative TonB-dependent receptor [PA1922] | - | 113 | 2 | CCG | P | P | P | P | P | P | P | L | L | L | L | L | P | P | P | P | P | P | P | P | P | P |
| 3884310 G | OPLDDDIH_03557 | conserved hypothetical protein [PA1854] | + | 937 | 1 | GGG | G | G | G | R | G | G | G | G | G | G | G | G | G | G | G | G | G | G | G | G | G | G |
| 3893909 T | OPLDDDIH_03567 | hypothetical protein [PA1844] | + | 46 | 1 | TGG | W | W | W | W | W | R | W | W | W | W | W | W | W | W | W | W | W | W | W | W | W | W |
| 3903027 G | OPLDDDIH_03575 | sulfite reductase [PA1838] | + | 1633 | 1 | GAG | E | E | E | K | E | E | E | K | K | K | K | K | E | K | K | K | E | K | X | E | K | E |
| 3927602 A | OPLDDDIH_03601 | membrane-bound lytic murein transglycosylase D precursor [PA1812] | + | 641 | 2 | TAC | Y | Y | Y | Y | Y | C | Y | Y | Y | Y | Y | Y | Y | Y | Y | Y | Y | Y | Y | Y | Y | Y |
| 3959542 T | OPLDDDIH_03631 | putative bacteriophage protein [PA0638] | - | 472 | 1 | ATG | M | M | M | M | M | M | M | V | V | V | V | V | M | M | M | M | M | M | M | M | M | M |
| 4015912 G | OPLDDDIH_03713 | hypothetical protein [PA1789] | + | 380 | 2 | CGG | R | R | R | Q | R | Q | Q | Q | Q | Q | Q | Q | R | Q | X | Q | R | Q | Q | R | Q | R |
| 4016044 T | OPLDDDIH_03713 | hypothetical protein [PA1789] | + | 512 | 2 | GTG | V | V | V | A | V | V | V | V | V | V | V | V | V | V | V | V | V | V | V | V | V | V |
| 4024311 G | OPLDDDIH_03719 | nitrate transporter [PA1783] | + | 1142 | 2 | GGC | G | G | G | D | G | D | D | D | D | D | D | D | G | D | D | D | G | D | D | G | D | G |
| 4036481 G | OPLDDDIH_03729 | CmaX protein [PA1773] | - | 190 | 1 | CTC | L | L | L | L | L | L | L | L | L | L | F | L | L | L | L | L | L | L | L | L | L | L |
| 4038847 A | OPLDDDIH_03732 | phosphoenolpyruvate synthase [PA1770] | - | 2062 | 1 | TTC | F | F | F | F | F | F | F | L | L | L | L | L | F | L | X | L | F | L | L | F | L | F |
| 4082293 T | OPLDDDIH_03773 | conserved hypothetical protein [PA1730] | - | 184 | 1 | ACC | T | T | T | T | T | A | T | T | T | T | T | T | T | T | T | T | T | T | T | T | T | T |
| 4115657 T | OPLDDDIH_03814 | conserved hypothetical protein [PA1689] | - | 1145 | 2 | TAC | Y | Y | Y | Y | Y | Y | Y | Y | Y | Y | Y | Y | Y | C | Y | Y | Y | Y | Y | Y | Y | Y |
| 4130627 A | OPLDDDIH_03832 | hypothetical protein [PA1672] | + | 289 | 1 | ACG | T | T | T | T | T | A | T | T | T | T | T | T | T | T | T | T | T | T | T | T | T | T |
| 4143551 G | OPLDDDIH_03842 | putative ClpA/B-type protease [PA1662] | - | 755 | 2 | GCC | A | A | A | V | A | V | V | V | V | V | V | V | A | X | V | V | A | V | X | A | V | A |
| 4178541 C | OPLDDDIH_03871 | hypothetical protein [PA1639] | - | 71 | 2 | GGC | G | G | G | G | G | G | D | G | G | G | G | G | G | G | G | G | G | G | G | G | G | G |
| 4216441 G | OPLDDDIH_03906 | hypothetical protein [PA1604] | + | 303 | 3 | TGG | W | W | W | W | W | W | W | W | W | W | W | W | W | W | W | W | W | W | W | W | * | W |
| 4231618 T | OPLDDDIH_03921 | succinyl-CoA synthetase alpha chain [PA1589] | - | 772 | 1 | ACC | T | T | T | T | T | T | T | T | T | T | T | T | T | A | A | T | T | T | T | T | T | T |
| 4232627 T | OPLDDDIH_03922 | succinyl-CoA synthetase beta chain [PA1588] | - | 929 | 2 | AAC | N | N | N | N | N | N | N | N | N | N | N | N | N | N | N | N | N | N | N | N | S | N |
| 4297084 C | OPLDDDIH_03983 | DNA ligase [PA1529] | - | 628 | 1 | GCC | A | A | A | T | A | T | T | T | T | T | T | T | A | T | T | T | A | X | X | A | T | A |
| 4317518 T | OPLDDDIH_03999 | hypothetical protein [PA1513] | + | 272 | 2 | GTG | V | V | V | A | V | V | V | V | V | V | V | V | V | V | V | V | V | V | V | V | V | V |
| 4325707 T | OPLDDDIH_04005 | hypothetical protein [PA1509] | + | 395 | 2 | GTG | V | V | V | V | V | A | V | V | V | V | V | V | V | V | V | V | V | V | V | V | V | V |
| 4327904 G | OPLDDDIH_04007 | putative transporter [PA1507] | + | 494 | 2 | GGC | G | G | G | G | G | G | G | D | G | G | G | G | G | G | G | G | G | G | G | G | G | G |
| 4336750 A | OPLDDDIH_04015 | conserved hypothetical protein [PA1499] | + | 407 | 2 | GAC | D | D | D | D | D | D | D | D | D | D | D | D | D | G | G | D | D | D | D | D | D | D |
| 4337277 G | OPLDDDIH_04015 | conserved hypothetical protein [PA1499] | + | 934 | 1 | GGC | G | G | G | G | G | G | G | S | S | S | S | S | G | G | G | G | G | G | G | G | G | G |
| 4341763 T | OPLDDDIH_04019 | hypothetical protein [PA1495] | + | 536 | 2 | CTG | L | L | L | L | L | L | L | L | L | L | L | L | L | L | L | L | L | L | X | L | P | L |
| 4354883 G | OPLDDDIH_04030 | putative transcriptional regulator [PA1484] | + | 323 | 2 | CGG | R | R | R | R | R | R | R | R | R | R | R | R | R | R | R | R | R | R | Q | R | R | R |
| 4386268 G | OPLDDDIH_04064 | conserved hypothetical protein [PA1450] | - | 701 | 2 | ACG | T | T | T | T | T | T | T | T | T | T | T | T | T | M | M | M | T | T | X | T | T | T |
| 4398297 T | OPLDDDIH_04078 | putative Resistance-Nodulation-Cell Division (RND) efflux transporter [PA1436] | - | 2249 | 2 | TAC | Y | Y | Y | C | Y | Y | Y | C | C | C | C | C | Y | C | C | C | Y | C | X | Y | C | Y |
| 4405750 T | OPLDDDIH_04084 | transcriptional regulator LasR [PA1430] | - | 591 | 3 | ATA | I | I | I | M | I | I | I | I | I | I | I | I | I | I | I | I | I | I | I | I | I | I |
| 4405998 T | OPLDDDIH_04084 | transcriptional regulator LasR [PA1430] | - | 343 | 1 | ACC | T | T | T | T | T | T | A | T | T | T | T | T | T | T | T | T | T | T | T | T | T | T |
| 4406061 G | OPLDDDIH_04084 | transcriptional regulator LasR [PA1430] | - | 280 | 1 | CAG | Q | Q | Q | Q | Q | Q | Q | Q | Q | Q | Q | Q | Q | Q | Q | Q | * | Q | Q | * | Q | * |
| 4406267 A | OPLDDDIH_04084 | transcriptional regulator LasR [PA1430] | - | 74 | 2 | ATG | M | K | K | K | K | K | K | K | K | K | K | K | K | K | K | K | K | K | K | K | K | K |
| 4418972 G | OPLDDDIH_04095 | putative transporter [PA1419] | + | 280 | 1 | GTC | V | V | V | I | V | I | I | I | I | I | I | I | V | I | I | I | V | X | I | V | I | V |
| 4432553 G | OPLDDDIH_04106 | hypothetical protein [PA1408] | + | 1081 | 1 | GGC | G | G | G | G | G | G | G | G | G | G | G | G | G | G | G | G | G | G | G | G | S | G |
| 4460832 G | OPLDDDIH_04132 | isocitrate dehydrogenase kinase/phosphatase [PA1376] | + | 997 | 1 | GGC | G | G | G | G | G | G | G | G | G | G | G | G | G | G | G | S | G | G | G | G | G | G |
| 4469160 T | OPLDDDIH_04137 | putative siderophore receptor [PA1365] | - | 1532 | 2 | CAG | Q | Q | Q | R | Q | R | R | R | R | R | R | R | Q | R | X | R | Q | R | R | X | R | Q |
| 4479921 G | OPLDDDIH_04150 | conserved hypothetical protein [PA1352] | + | 88 | 1 | GCC | A | A | A | A | A | A | A | A | A | A | A | T | A | A | A | A | A | A | A | A | A | A |
| 4488126 T | OPLDDDIH_04158 | hypothetical protein [PA1345] | - | 322 | 1 | AGC | S | S | S | S | S | S | S | S | S | S | S | S | S | S | S | S | S | S | S | S | G | S |
| 4491708 T | OPLDDDIH_04162 | amino acid ABC transporter membrane protein [PA1341] | + | 167 | 2 | GTG | V | V | V | V | V | V | V | V | V | V | V | V | V | A | V | V | V | V | V | V | V | V |
| 4500651 G | OPLDDDIH_04169 | putative oxidoreductase [PA1334] | + | 388 | 1 | GTC | V | V | V | V | V | V | V | V | V | I | V | V | V | V | X | V | V | V | V | V | V | V |
| 4504789 T | OPLDDDIH_04173 | putative short-chain dehydrogenase [PA1330] | - | 326 | 2 | AAC | N | N | N | N | N | N | N | N | N | N | S | N | N | N | N | N | N | N | N | N | N | N |
| 4522947 G | OPLDDDIH_04190 | putative major facilitator superfamily (MFS) transporter [PA1313] | - | 857 | 2 | CCC | P | P | P | P | P | L | P | P | P | P | P | P | P | P | P | P | P | P | P | P | P | P |
| 4554128 T | OPLDDDIH_04221 | putative major facilitator superfamily (MFS) transporter [PA1282] | + | 1048 | 1 | TCC | S | S | S | S | S | S | S | P | P | P | P | P | S | P | P | P | S | X | X | S | P | S |
| 4556501 G | OPLDDDIH_04224 | nicotinate-nucleotide--dimethylbenzimidazole phosphoribosyltransferase [PA1279] | - | 425 | 2 | GCG | A | A | A | A | A | A | V | A | A | A | A | A | A | A | A | A | A | A | A | X | A | A |
| 4566560 A | OPLDDDIH_04233 | hypothetical protein [PA1270] | - | 1187 | 2 | GTG | V | V | V | V | V | V | A | V | V | V | V | V | V | V | V | V | V | V | V | V | V | V |
| 4573621 G | OPLDDDIH_04240 | putative transcriptional regulator [PA1264] | + | 329 | 2 | CGT | R | R | R | R | R | H | R | R | R | R | R | R | R | R | R | R | R | R | R | R | R | R |
| 4582956 G | OPLDDDIH_04249 | hypothetical protein [PA1255] | + | 292 | 1 | GCC | A | A | A | A | A | A | A | T | A | A | A | A | A | A | A | A | A | A | A | A | A | A |
| 4597552 T | OPLDDDIH_04259 | hypothetical protein [PA1245] | - | 170 | 2 | GAC | D | D | D | D | D | D | D | D | D | G | D | D | D | D | D | D | D | D | D | D | D | D |
| 4613859 T | OPLDDDIH_04271 | hypothetical protein [PA1242] | - | 704 | 2 | GAC | D | D | D | D | D | D | D | D | D | D | D | G | D | D | D | D | D | X | D | D | D | D |
| 4621151 C | OPLDDDIH_04278 | putative major facilitator superfamily (MFS) transporter [PA1236] | + | 1078 | 1 | CGC | R | R | R | R | R | R | C | R | R | R | R | R | R | R | R | R | R | R | R | R | R | R |
| 4626726 A | OPLDDDIH_04285 | putative transcriptional regulator [PA1229] | + | 95 | 2 | AAC | N | N | N | N | N | N | N | N | N | N | N | N | N | N | N | N | N | S | N | N | N | N |
| 4636932 T | OPLDDDIH_04294 | hypothetical protein [PA1220] | + | 1177 | 1 | TGG | W | W | W | W | W | R | W | W | W | W | W | W | W | W | W | W | W | W | W | X | W | W |
| 4716608 T | OPLDDDIH_04374 | putative transcriptional regulator [PA1182] | + | 173 | 2 | CTC | L | L | L | L | L | P | L | L | L | L | L | L | L | L | L | L | L | L | L | L | L | L |
| 4741496 T | OPLDDDIH_04395 | rRNA methyltransferase [PA1161] | + | 178 | 1 | TTC | F | F | F | L | F | F | F | F | F | F | F | F | F | F | F | F | F | F | F | F | F | F |
| 4762346 G | OPLDDDIH_04408 | putative amino acid permease [PA1147] | + | 781 | 1 | GTG | V | V | V | V | V | V | V | V | V | V | V | V | V | V | V | V | V | V | V | V | M | V |
| 4768940 G | OPLDDDIH_04414 | putative transcriptional regulator [PA1141] | + | 182 | 2 | CGC | R | R | R | R | R | R | R | R | R | R | R | R | R | R | R | H | R | R | R | R | R | R |
| 4770152 T | OPLDDDIH_04415 | conserved hypothetical protein [PA1140] | + | 355 | 1 | TTC | F | F | F | F | F | F | F | F | F | F | F | F | F | F | F | F | F | F | F | F | L | F |
| 4772371 G | OPLDDDIH_04417 | putative transcriptional regulator [PA1138] | - | 176 | 2 | CCG | P | P | P | L | P | P | P | L | L | L | L | L | P | L | L | L | P | L | L | P | L | P |
| 4787414 G | OPLDDDIH_04435 | diguanylate cyclase TpbB [PA1120] | + | 26 | 2 | GGT | G | G | G | G | G | G | G | G | G | G | G | D | G | G | G | G | G | G | G | G | G | G |
| 4825319 T | OPLDDDIH_04471 | Carnitine monooxygenase oxygenase subunit | - | 320 | 2 | AAC | N | N | N | S | N | N | N | N | N | N | N | N | N | N | N | N | N | N | N | N | N | N |
| 4828986 T | OPLDDDIH_04476 | ArnB [PA3552] | - | 565 | 1 | ACC | T | T | T | T | T | A | T | T | T | T | T | T | T | T | T | T | T | T | T | T | T | T |
| 4832327 G | OPLDDDIH_04478 | flagellar hook-associated protein 1 FlgK [PA1086] | - | 901 | 1 | CTC | L | L | L | L | L | F | L | L | L | L | L | L | L | L | L | L | L | L | L | L | L | L |
| 4856788 T | OPLDDDIH_04504 | hypothetical protein [PA1060] | - | 676 | 1 | ACG | T | T | T | T | T | A | T | T | T | T | T | T | T | T | T | T | T | T | T | T | T | T |
| 4874565 T | OPLDDDIH_04520 | hypothetical protein [PA1045] | - | 1378 | 1 | AGC | S | S | S | G | S | G | G | G | G | G | G | G | S | G | G | G | S | X | G | S | G | S |
| 4884570 T | OPLDDDIH_04533 | QuiP [PA1032] | + | 1396 | 1 | TGG | W | W | W | W | W | W | W | W | W | W | R | W | W | W | W | W | W | W | W | W | W | W |
| 4891866 A | OPLDDDIH_04538 | putative aldehyde dehydrogenase [PA1027] | - | 305 | 2 | GTC | V | V | V | V | V | V | V | V | V | V | V | V | V | A | A | V | V | V | V | V | V | V |
| 4919545 T | OPLDDDIH_04567 | anthranilate synthase component II [PA1002] | - | 322 | 1 | AGC | S | S | S | S | S | S | S | S | S | S | S | S | S | S | S | G | S | S | S | S | S | S |
| 4936523 T | OPLDDDIH_04583 | hypothetical protein [PA0988] | - | 355 | 1 | ACC | T | T | T | T | T | T | T | T | T | T | T | T | T | T | T | A | T | T | T | T | T | T |
| 4952409 G | OPLDDDIH_04596 | glutathione-regulated potassium-efflux system protein KefB [PA1207] | - | 701 | 2 | ACC | T | T | T | T | T | T | T | T | T | I | T | T | T | T | T | T | T | T | T | T | T | T |
| 4956162 G | OPLDDDIH_04600 | betaine aldehyde dehydrogenase [PA5373] | - | 1136 | 2 | CCC | P | P | P | P | P | P | P | L | L | L | L | L | P | P | P | P | P | P | P | P | P | P |
| 4960993 G | OPLDDDIH_04603 | putative dehydrogenase [PA4100] | + | 1619 | 2 | GGC | G | G | G | D | G | G | G | G | G | G | G | G | G | G | G | G | G | X | G | G | G | G |
| 4962394 G | OPLDDDIH_04605 | gentisate 1,2-dioxygenase [PA2470] | + | 344 | 2 | GGC | G | G | G | G | G | G | G | G | G | G | G | G | G | G | G | D | G | G | G | G | G | G |
| 4967864 A | OPLDDDIH_04608 | Tyrosine recombinase XerC | + | 862 | 1 | ACC | T | T | T | A | T | T | T | A | A | A | A | A | T | A | A | A | T | A | A | T | A | T |
| 4976008 T | OPLDDDIH_04619 | Holliday junction DNA helicase RuvB [PA0967] | - | 791 | 2 | CAC | H | H | H | H | H | H | H | H | H | H | H | H | H | H | H | H | H | R | H | H | H | H |
| 4977831 T | OPLDDDIH_04621 | Holliday junction resolvase RuvC [PA0965] | - | 125 | 2 | CAC | H | H | H | H | H | H | H | H | H | H | H | H | H | H | H | H | H | H | H | H | R | H |
| 4978905 T | OPLDDDIH_04623 | aspartyl-tRNA synthetase [PA0963] | - | 1724 | 2 | AAG | K | K | K | K | K | K | K | R | K | K | K | K | K | K | K | K | K | K | K | K | K | K |
| 4984788 G | OPLDDDIH_04628 | Basic amino acid, basic peptide and imipenem outer membrane porin OprD precursor [PA0958] | + | 1017 | 3 | TGG | W | W | * | * | * | * | * | * | * | * | * | * | * | * | * | * | * | * | * | * | * | * |
| 4990659 C | OPLDDDIH_04636 | putative ribonuclease [PA0951] | - | 778 | 1 | GCG | A | A | A | T | A | A | A | A | A | A | A | A | A | A | A | A | A | A | A | A | A | A |
| 5001299 A | OPLDDDIH_04651 | conserved hypothetical protein [PA0937] | - | 371 | 2 | CTG | L | L | L | L | L | P | L | L | L | L | L | L | L | L | L | L | L | L | L | L | L | L |
| 5003907 A | OPLDDDIH_04654 | GTP pyrophosphokinase [PA0934] | - | 2156 | 2 | ATT | I | I | I | T | I | I | I | T | T | T | T | T | I | T | T | T | I | T | T | I | T | I |
| 5019483 C | OPLDDDIH_04664 | hypothetical protein [PA0924] | + | 1303 | 1 | CAG | Q | Q | Q | Q | Q | Q | Q | Q | Q | Q | Q | Q | Q | * | * | Q | Q | Q | Q | Q | Q | Q |
| 5046671 G | OPLDDDIH_04694 | succinylglutamate 5-semialdehyde dehydrogenase [PA0898] | - | 1054 | 1 | CCG | P | P | P | P | P | P | P | S | S | S | S | S | P | P | P | P | P | P | P | P | P | P |
| 5067270 T | OPLDDDIH_04713 | putative acyl-CoA dehydrogenase [PA0879] | - | 511 | 1 | ACG | T | T | T | T | T | T | T | T | T | T | A | T | T | T | T | T | T | T | T | T | T | T |
| 5077001 T | OPLDDDIH_04722 | aromatic amino acid aminotransferase [PA0870] | + | 112 | 1 | TAC | Y | Y | Y | Y | Y | Y | H | Y | Y | Y | Y | Y | Y | Y | Y | Y | Y | Y | Y | Y | Y | Y |
| 5086502 G | OPLDDDIH_04732 | hypothetical protein [PA0861] | - | 1334 | 2 | CCG | P | P | P | P | P | P | P | L | P | P | P | P | P | P | P | P | P | P | P | P | P | P |
| 5094659 T | OPLDDDIH_04739 | hypothetical protein [PA0855] | + | 869 | 2 | CTG | L | L | L | L | L | L | L | P | P | P | P | P | L | L | L | L | L | L | X | X | L | L |
| 5120982 T | OPLDDDIH_04761 | hypothetical protein [PA0833] | - | 478 | 1 | ACC | T | T | T | T | T | T | T | A | A | A | A | A | T | T | T | T | T | T | T | T | T | T |
| 5124181 T | OPLDDDIH_04764 | hypothetical protein [PA0830] | + | 730 | 1 | TAC | Y | Y | Y | H | Y | H | H | H | H | H | H | H | Y | H | H | H | Y | H | H | Y | H | Y |
| 5165678 A | OPLDDDIH_04824 | hypothetical protein | + | 341 | 2 | CAC | H | H | H | R | H | R | R | R | R | R | R | R | H | R | R | R | H | R | R | H | R | H |
| 5169635 T | OPLDDDIH_04828 | hypothetical protein [PA0826] | + | 451 | 1 | TTC | F | F | F | F | F | F | F | F | F | F | F | F | F | F | F | L | F | F | F | F | F | F |
| 5202810 A | OPLDDDIH_04860 | propionate catabolic protein PrpD [PA0792] | + | 1276 | 1 | AGC | S | S | S | S | S | S | S | S | S | S | S | S | S | S | S | S | S | S | G | S | S | S |
| 5203464 G | OPLDDDIH_04861 | putative transcriptional regulator [PA0791] | - | 322 | 1 | CGG | R | R | R | W | R | W | W | W | W | W | W | W | R | W | W | W | R | W | W | R | W | R |
| 5213638 G | OPLDDDIH_04870 | putative transcriptional regulator [PA0784] | + | 593 | 2 | GGC | G | G | G | G | G | G | G | G | G | G | G | G | G | G | G | G | G | G | D | G | G | G |
| 5217659 T | OPLDDDIH_04872 | proline dehydrogenase PutA [PA0782] | - | 1463 | 2 | AAC | N | N | N | N | N | N | N | N | S | N | N | N | N | N | N | N | N | N | N | N | N | N |
| 5220792 T | OPLDDDIH_04873 | hypothetical protein [PA0781] | + | 1208 | 2 | GTG | V | V | V | V | V | V | A | V | V | V | V | V | V | V | V | V | V | V | V | V | V | V |
| 5249675 T | OPLDDDIH_04903 | conserved hypothetical protein [PA0752] | + | 140 | 2 | CTG | L | L | L | L | L | L | L | P | P | P | P | P | L | L | L | L | L | L | X | L | L | L |
| 5265923 T | OPLDDDIH_04919 | hypothetical protein [PA0737] | - | 326 | 2 | AAC | N | N | N | N | N | N | N | N | N | N | N | N | N | S | N | N | N | N | N | N | N | N |
| 5266444 G | OPLDDDIH_04920 | hypothetical protein | - | 64 | 1 | CCG | P | P | P | P | P | A | P | P | P | P | P | P | P | P | P | P | P | P | P | P | P | P |
| 5270361 G | OPLDDDIH_04925 | hypothetical protein [PA0732] | + | 607 | 1 | GCC | A | A | A | A | A | A | A | A | A | A | A | A | A | A | A | A | A | T | A | A | A | A |
| 5276619 G | OPLDDDIH_04935 | putative transcriptional regulator [PA0708] | + | 250 | 1 | GGC | G | G | G | G | G | G | G | S | S | S | S | S | G | G | G | G | G | G | G | G | G | G |
| 5306581 T | OPLDDDIH_04955 | hypothetical protein [PA0690] | - | 2594 | 2 | CAG | Q | Q | Q | Q | Q | Q | R | Q | Q | Q | Q | Q | Q | Q | Q | Q | Q | Q | Q | Q | Q | Q |
| 5317190 T | OPLDDDIH_04961 | hypothetical protein | - | 695 | 2 | AAC | N | N | N | N | N | S | N | N | N | N | N | N | N | N | N | N | N | N | N | N | N | N |
| 5319961 G | OPLDDDIH_04965 | hypothetical protein [PA0679] | - | 385 | 1 | CAG | Q | Q | Q | Q | Q | Q | Q | Q | Q | Q | Q | Q | Q | Q | Q | Q | Q | Q | * | Q | Q | Q |
| 5337505 C | OPLDDDIH_04978 | exonuclease SbcD [PA4281] | + | 908 | 2 | GCC | A | A | A | A | A | A | V | A | A | A | A | A | A | A | A | A | A | A | A | A | A | A |
| 5338503 T | OPLDDDIH_04979 | putative exonuclease [PA4282] | + | 668 | 2 | CTG | L | L | L | P | L | L | L | L | L | L | L | L | L | L | L | L | L | L | L | L | L | L |
| 5365788 C | OPLDDDIH_04996 | TadD [PA4299] | - | 527 | 2 | GGC | G | G | G | G | G | G | G | G | G | G | G | G | G | G | G | D | G | G | G | G | G | G |
| 5369749 C | OPLDDDIH_05000 | TadZ [PA4303] | - | 811 | 1 | GCC | A | A | A | T | A | T | T | T | T | T | T | T | A | T | X | T | A | T | T | A | T | A |
| 5389662 G | OPLDDDIH_05016 | hypothetical protein [PA4318] | + | 719 | 2 | GGC | G | G | G | D | G | D | D | D | D | D | D | D | G | D | D | D | G | D | X | G | D | G |
| 5406882 G | OPLDDDIH_05032 | putative transport protein [PA4334] | - | 11 | 2 | TCC | S | S | S | S | S | S | S | F | F | F | F | F | S | S | S | S | S | S | S | S | S | S |
| 5415251 G | OPLDDDIH_05041 | putative major facilitator superfamily (MFS) transporter [PA4343] | - | 281 | 2 | ACG | T | T | T | T | T | T | T | T | T | T | T | T | T | T | T | T | T | M | T | T | T | T |
| 5484901 T | OPLDDDIH_05109 | cell division protein FtsQ [PA4409] | - | 79 | 1 | AGC | S | S | S | S | S | S | S | G | S | S | S | S | S | S | S | S | S | S | S | S | S | S |

| | | | | | | | | | | | | | | | | | | | | | | | | | | | | | |
|---|---|---|---|---|---|---|---|---|---|---|---|---|---|---|---|---|---|---|---|---|---|---|---|---|---|---|---|---|---|
| 5495139 | C | OPLDDDIH_05118 | penicillin-binding protein 3 [PA4418] | - | 1511 | 2 | CGC | R | R | R | H | R | H | H | H | H | H | H | H | R | H | H | H | R | H | H | R | H | R |
| 5495550 | T | OPLDDDIH_05118 | penicillin-binding protein 3 [PA4418] | - | 1100 | 2 | TAC | Y | Y | Y | Y | Y | Y | Y | C | C | C | C | C | Y | Y | Y | Y | Y | Y | Y | Y | Y | Y |
| 5508808 | T | OPLDDDIH_05134 | putative oxidoreductase [PA4434] | + | 313 | 1 | TTC | F | F | F | F | F | F | F | F | F | F | F | F | F | L | L | L | F | F | L | F | F | F |
| 5552131 | T | OPLDDDIH_05177 | cytoplasmic axial filament protein [PA4477] | - | 275 | 2 | CAG | Q | Q | Q | Q | Q | Q | Q | Q | Q | R | Q | Q | Q | Q | Q | Q | Q | Q | Q | Q | Q | Q |
| 5589190 | G | OPLDDDIH_05208 | putative transcriptional regulator [PA4508] | + | 88 | 1 | GGC | G | G | G | S | G | G | G | G | G | G | G | G | G | G | G | G | G | G | G | G | G | G |
| 5592822 | A | OPLDDDIH_05212 | lipopolysaccharide biosynthetic protein LpxO1 [PA4512] | + | 757 | 1 | ACC | T | T | T | T | T | A | T | T | T | T | T | T | T | T | T | T | T | T | T | T | T | T |
| 5624154 | G | OPLDDDIH_05238 | type 4 fimbrial biogenesis protein PilB [PA4526] | + | 1072 | 1 | GTG | V | V | V | M | V | M | M | M | M | M | M | M | V | M | M | M | V | M | M | V | M | V |
| 5640035 | T | OPLDDDIH_05253 | hypothetical protein [PA4541] | + | 4279 | 1 | TGA | * | * | * | * | * | * | * | * | * | * | * | * | * | * | * | * | * | R | * | * | * | * |
| 5640750 | T | OPLDDDIH_05254 | chromosome partitioning protein Soj [PA5563] | + | 346 | 1 | TCC | S | S | S | P | S | S | S | P | P | P | P | P | S | P | P | P | S | P | P | S | P | S |
| 5675247 | G | OPLDDDIH_05292 | Toxin coregulated pilus biosynthesis protein E | + | 939 | 3 | TGG | W | W | W | W | W | * | W | W | W | W | W | W | W | W | W | W | W | W | W | W | W | W |
| 5716660 | A | OPLDDDIH_05343 | hypothetical protein | + | 412 | 1 | ACC | T | T | T | T | T | T | T | T | T | T | T | T | T | T | T | T | T | T | T | T | A | T |
| 5762812 | G | OPLDDDIH_05387 | GTP-binding protein Obg [PA4566] | - | 955 | 1 | CGC | R | R | R | R | R | R | R | C | R | R | R | R | R | R | R | R | R | R | R | R | R | R |
| 5773712 | T | OPLDDDIH_05398 | hypothetical protein [PA4577] | - | 95 | 2 | CAT | H | H | H | H | H | H | R | H | H | H | H | H | H | H | H | H | H | H | H | H | H | H |
| 5794868 | T | OPLDDDIH_05417 | putative ATP-binding component of ABC transporter [PA4595] | - | 1580 | 2 | TAC | Y | Y | Y | Y | Y | C | Y | Y | Y | Y | Y | Y | Y | Y | Y | Y | Y | Y | Y | Y | Y | Y |
| 5794949 | T | OPLDDDIH_05417 | putative ATP-binding component of ABC transporter [PA4595] | - | 1499 | 2 | GAC | D | D | D | D | D | D | D | D | D | D | D | D | D | D | D | D | D | D | D | D | G | D |
| 5816170 | G | OPLDDDIH_05432 | DNA repair protein RadA [PA4609] | - | 353 | 2 | CCG | P | P | P | P | P | P | P | P | P | P | P | P | P | P | P | P | P | L | P | P | P | P |
| 5839474 | T | OPLDDDIH_05449 | hypothetical protein [PA4625] | - | 1283 | 2 | TAC | Y | Y | Y | Y | Y | C | Y | Y | Y | Y | Y | Y | Y | Y | Y | Y | Y | Y | Y | Y | Y | Y |
| 5843359 | G | OPLDDDIH_05452 | lysine-specific permease [PA4628] | + | 109 | 1 | GCC | A | A | A | A | A | A | A | A | A | A | A | A | A | A | A | T | A | A | A | A | A | A |
| 5905690 | G | OPLDDDIH_05515 | ferric iron-binding periplasmic protein HitA [PA4687] | + | 163 | 1 | GGC | G | G | G | S | G | S | S | S | S | S | S | S | G | S | S | S | G | S | S | G | S | G |
| 5950537 | A | OPLDDDIH_05554 | suppressor protein DksA [PA4723] | + | 323 | 2 | TAC | Y | Y | Y | Y | Y | Y | Y | C | C | C | C | C | Y | Y | Y | Y | Y | Y | Y | Y | Y | Y |
| 5967393 | T | OPLDDDIH_05567 | hypothetical protein [PA4735] | + | 991 | 1 | TTC | F | F | F | F | F | F | F | F | F | F | F | F | F | F | F | F | F | F | F | F | L | F |
| 6060605 | G | OPLDDDIH_05664 | 2,4-dienoyl-CoA reductase FadH2 [PA4814] | + | 31 | 1 | GCT | A | A | A | A | A | A | A | A | A | A | A | A | A | A | A | A | A | A | T | A | A | A |
| 6075761 | G | OPLDDDIH_05677 | hypothetical protein [PA4826] | - | 80 | 2 | CCC | P | P | P | P | P | P | P | P | P | P | P | P | P | P | P | P | P | P | P | P | L | P |
| 6099184 | G | OPLDDDIH_05699 | 3-dehydroquinate dehydratase [PA4846] | + | 62 | 2 | GGC | G | G | G | G | G | G | D | G | G | G | G | G | G | G | G | G | G | G | G | G | G | G |
| 6103214 | G | OPLDDDIH_05703 | ribosomal protein L11 methyltransferase [PA4850] | + | 730 | 1 | GGC | G | G | G | G | G | G | G | S | S | S | S | S | G | G | G | G | G | G | G | G | G | G |
| 6153830 | T | OPLDDDIH_05753 | putative aldehyde dehydrogenase [PA4899] | - | 1346 | 2 | AAC | N | N | N | N | N | N | S | N | N | N | N | N | N | N | N | N | N | N | N | N | N | N |
| 6159698 | G | OPLDDDIH_05757 | putative major facilitator superfamily (MFS) transporter [PA4903] | - | 860 | 2 | ACC | T | T | T | T | T | T | T | T | T | T | T | T | T | T | T | T | T | T | T | T | I | T |
| 6188265 | A | OPLDDDIH_05782 | conserved hypothetical protein [PA4928] | - | 530 | 2 | ATC | I | I | I | I | I | I | I | I | I | I | I | N | I | I | I | I | I | I | I | I | I | I |
| 6190693 | G | OPLDDDIH_05783 | hypothetical protein [PA4929] | + | 1687 | 1 | GCC | A | A | A | A | A | T | A | A | A | A | A | A | A | A | A | A | A | A | A | A | A | A |
| 6204161 | G | OPLDDDIH_05796 | conserved hypothetical protein [PA4939] | - | 62 | 2 | GCG | A | A | A | A | A | A | A | A | A | V | A | A | A | A | A | A | A | A | A | A | A | A |
| 6209885 | T | OPLDDDIH_05803 | DNA mismatch repair protein MutL [PA4946] | - | 1415 | 2 | CAC | H | H | H | L | H | L | L | L | L | L | L | L | H | L | L | L | H | L | L | H | L | H |
| 6232625 | G | OPLDDDIH_05821 | topoisomerase IV subunit A [PA4964] | - | 1480 | 1 | CGC | R | R | R | R | R | R | R | R | R | R | R | R | R | R | R | R | R | R | C | R | R | R |
| 6236160 | T | OPLDDDIH_05824 | topoisomerase IV subunit B [PA4967] | - | 1376 | 2 | GAG | E | E | E | E | E | E | E | E | E | E | E | E | E | E | E | E | E | E | E | E | G | E |
| 6242015 | T | OPLDDDIH_05830 | thiamin biosynthesis protein ThiC [PA4973] | - | 1250 | 2 | AAG | K | K | K | K | K | K | R | K | K | K | K | K | K | K | K | K | K | K | K | K | K | K |
| 6253364 | T | OPLDDDIH_05838 | putative amino acid permease [PA4981] | - | 1352 | 2 | TAC | Y | Y | Y | Y | Y | Y | Y | Y | Y | Y | Y | Y | Y | C | C | Y | Y | Y | Y | Y | Y | Y |
| 6282471 | T | OPLDDDIH_05862 | putative glycosyl transferase [PA5004] | - | 206 | 2 | GAC | D | D | D | G | D | D | D | G | G | G | G | G | D | G | G | G | D | G | X | D | G | D |
| 6285977 | G | OPLDDDIH_05864 | hypothetical protein [PA5006] | - | 23 | 2 | GCC | A | A | A | A | A | A | A | A | A | A | A | A | A | A | A | A | A | A | A | A | V | A |
| 6290909 | G | OPLDDDIH_05870 | heptosyltransferase II [PA5012] | - | 599 | 2 | GCC | A | A | A | A | A | V | A | A | A | A | A | A | A | A | A | A | A | A | A | A | A | A |
| 6326947 | G | OPLDDDIH_05893 | glutamate synthase small chain [PA5035] | - | 344 | 2 | GCC | A | A | A | A | A | A | A | V | V | V | V | V | A | A | A | A | A | A | A | A | A | A |
| 6336145 | T | OPLDDDIH_05898 | Type 4 fimbrial biogenesis outer membrane protein PilQ precursor [PA5040] | - | 1325 | 2 | GAC | D | D | G | D | G | D | D | D | D | D | D | D | D | D | D | D | D | D | D | D | D | D |
| 6367487 | T | OPLDDDIH_05928 | transport protein TatC [PA5070] | + | 683 | 2 | GTG | V | V | V | V | V | V | V | V | V | V | V | V | V | V | V | V | V | V | V | V | A | V |
| 6373292 | T | OPLDDDIH_05934 | putative binding protein component of ABC transporter [PA5076] | - | 310 | 1 | ACC | T | T | T | A | T | A | A | A | A | A | A | A | T | A | A | A | T | A | A | T | A | T |
| 6375077 | A | OPLDDDIH_05935 | periplasmic glucans biosynthesis protein MdoH [PA5077] | - | 1348 | 1 | TCC | S | S | S | S | S | S | S | S | S | S | S | S | S | S | S | P | S | S | S | S | S | S |
| 6429058 | G | OPLDDDIH_05980 | hypothetical protein [PA5121] | - | 2030 | 2 | ACC | T | T | T | T | T | T | T | T | T | T | T | T | T | T | T | T | T | I | T | T | T | T |
| 6433423 | T | OPLDDDIH_05983 | two-component sensor NtrB [PA5124] | + | 689 | 2 | ATC | I | I | I | I | I | T | I | I | I | I | I | I | I | I | I | I | I | I | I | I | I | I |
| 6450840 | T | OPLDDDIH_06001 | hypothetical protein | - | 184 | 1 | ATC | I | I | I | V | I | V | V | V | V | V | V | V | I | V | V | V | I | V | V | I | V | I |
| 6452599 | G | OPLDDDIH_06004 | hypothetical protein [PA5144] | + | 181 | 1 | GGT | G | G | G | G | G | G | S | G | G | G | G | G | G | G | G | G | G | G | G | G | G | G |
| 6476322 | T | OPLDDDIH_06022 | hypothetical protein | - | 1079 | 2 | GAC | D | D | D | D | D | G | D | D | D | D | D | D | D | D | D | D | D | D | D | D | D | D |
| 6479561 | G | OPLDDDIH_06024 | hypothetical protein | - | 161 | 2 | TCA | S | S | S | S | S | S | S | S | S | S | S | L | S | S | S | S | S | S | S | S | S | S |
| 6501516 | T | OPLDDDIH_06044 | putative two-component response regulator [PA5166] | + | 10 | 1 | TTC | F | F | F | F | F | F | F | F | F | F | F | F | F | F | F | F | F | L | F | F | F | F |
| 6548333 | G | OPLDDDIH_06086 | putative phosphate transporter [PA5207] | - | 644 | 2 | ACG | T | T | T | M | T | T | T | M | M | M | M | M | T | M | M | M | T | M | M | T | M | T |
| 6559581 | G | OPLDDDIH_06095 | putative permease of ABC iron transporter [PA5216] | - | 928 | 1 | CAG | Q | Q | Q | Q | Q | Q | Q | Q | Q | Q | Q | Q | Q | Q | Q | * | Q | Q | Q | X | Q | Q |
| 6567736 | T | OPLDDDIH_06103 | aminopeptidase P [PA5224] | - | 1268 | 2 | AAC | N | N | N | N | N | N | N | N | N | N | N | N | N | N | N | N | N | S | N | N | N | N |
| 6594534 | A | OPLDDDIH_06128 | hypothetical protein [PA5248] | - | 1811 | 2 | GTG | V | V | V | V | V | V | V | A | A | A | A | A | V | A | A | A | V | A | A | V | A | V |
| 6607536 | T | OPLDDDIH_06141 | porphobilinogen deaminase [PA5260] | - | 878 | 2 | GAC | D | D | D | G | D | G | G | G | G | G | G | G | D | G | G | G | D | G | G | D | G | D |
| 6666775 | T | OPLDDDIH_06192 | Na(+)/H(+) antiporter NhaA | - | 881 | 2 | CAC | H | H | H | H | H | H | H | R | R | R | R | R | H | H | H | H | H | H | H | H | H | H |
| 6691722 | G | OPLDDDIH_06210 | hypothetical protein | + | 1449 | 3 | TGG | W | W | W | W | W | W | W | * | * | * | * | * | W | W | W | W | W | W | W | W | W | W |
| 6705544 | G | OPLDDDIH_06220 | hypothetical protein [PA5294] | + | 419 | 2 | GGC | G | G | G | G | G | G | G | G | G | G | G | G | G | G | G | D | G | G | G | G | G | G |
| 6728727 | T | OPLDDDIH_06238 | putative aldehyde dehydrogenase [PA5312] | - | 550 | 1 | ACC | T | T | T | T | T | T | T | T | T | T | T | T | T | A | A | A | T | T | A | T | T | T |
| 6730613 | G | OPLDDDIH_06239 | putative pyridoxal-dependent aminotransferase [PA5313] | + | 1108 | 1 | GTC | V | V | V | V | V | V | V | I | I | I | I | I | V | V | V | V | V | V | V | V | V | V |
| 6737103 | T | OPLDDDIH_06247 | Phosphoglucosamine mutase | + | 575 | 2 | CTG | L | L | L | L | L | L | L | L | L | L | L | L | L | L | L | L | L | L | P | L | L | L |
| 6741024 | T | OPLDDDIH_06249 | putative transcriptional regulator [PA5324] | - | 149 | 2 | GAC | D | D | D | D | D | D | G | D | D | D | D | D | D | D | D | D | D | D | D | D | D | D |
| 6779297 | T | OPLDDDIH_06291 | ATP-binding component of ABC phosphate transporter [PA5366] | - | 110 | 2 | AAC | N | N | N | N | N | N | N | N | N | N | N | N | N | N | N | N | N | N | S | N | N | N |
| 6831080 | T | OPLDDDIH_06338 | GbcA [PA5410] | - | 971 | 2 | CAC | H | H | H | H | H | H | H | H | H | H | H | H | H | H | H | H | H | R | H | H | H | H |
| 6855152 | G | OPLDDDIH_06357 | aspartate ammonia-lyase [PA5429] | + | 520 | 1 | GCC | A | A | A | A | A | A | A | A | A | A | A | A | A | A | A | T | A | A | A | A | A | A |
| 6856939 | G | OPLDDDIH_06358 | hypothetical protein [PA5430] | - | 698 | 2 | CCG | P | P | P | P | P | P | L | P | P | P | P | P | P | P | P | P | P | P | X | P | P | P |
| 6893484 | T | OPLDDDIH_06386 | hypothetical protein [PA5456] | + | 1090 | 1 | TAC | Y | Y | Y | Y | Y | Y | Y | Y | Y | Y | Y | Y | Y | Y | Y | Y | Y | Y | H | Y | Y | Y |
| 6897243 | A | OPLDDDIH_06391 | hypothetical protein [PA5461] | + | 248 | 2 | CAC | H | H | H | R | H | H | H | R | R | R | R | R | H | R | R | R | H | R | R | H | R | H |
| 6956482 | G | OPLDDDIH_06447 | putative potassium efflux transporter [PA5518] | - | 770 | 2 | GCC | A | A | A | A | A | A | A | A | A | A | A | A | A | A | A | A | A | A | V | A | A | A |
| 6971059 | G | OPLDDDIH_06460 | TonB1 [PA5531] | - | 349 | 1 | CTT | L | L | L | L | L | L | L | L | L | L | L | F | L | L | L | L | L | X | X | L | L | L |
| 6972414 | T | OPLDDDIH_06461 | Zinc-binding GTPase YeiR | - | 55 | 1 | ACC | T | T | T | T | T | T | A | T | T | T | T | T | T | T | T | T | T | T | T | T | T | T |
| 6977644 | A | OPLDDDIH_06468 | hypothetical protein [PA5539] | + | 494 | 2 | GAC | D | D | D | D | D | D | D | D | D | D | D | D | D | D | D | D | D | G | D | D | D | D |
| 6994274 | G | OPLDDDIH_06483 | ATP synthase beta chain [PA5554] | - | 881 | 2 | GCC | A | A | A | A | A | A | A | A | A | A | A | A | A | A | A | A | A | A | A | A | V | A |
